# Race- and sex-specific disease trajectory in individuals with rare variants in seven cerebrovascular small vessel disease genes: a genotype-first population study

**DOI:** 10.1101/2022.08.11.22278284

**Authors:** Jiang Li, Vida Abedi, Durgesh Chaudhary, Regeneron Genetics Center, Oded Goren, David J. Carey, Karen E. Wain, Ramin Zand

## Abstract

**Background:** This study is aimed to explore the risk for a lifetime or early onset of cardiovascular diseases and diseases within or beyond the circulatory system observed in rare variant(RV) carriers of seven well-characterized monogenic cerebral small vessel disease (CSVD) risk genes (*COL4A1, COL4A2, NOTCH3, HTRA1, TREX1, CTC1*, and *GLA*) using a genotype-first approach within a hospital system population-based biobank cohort.

**Method:** This was a retrospective longitudinal study. MyCode participants with sequenced exomes were temporally split into discovery (n=92,445) and replication (n=81,130) cohorts. A workflow was created to prioritize potentially pathogenic variants by integrating three variant annotation pipelines. After propensity score matching, the number of participants in both discovery and replication cohorts was 2738/5490 and 1695/3410 for carriers/noncarriers, respectively.

**Result:** Most of the RVs identified were in a heterozygous form and disproportionately present in participants with African ancestry. Carriers showed an increased risk for early signs and symptoms of cerebrovascular disease. Cox regression model showed *NOTCH3* (European), *TREX*1 (European), and COL4A1/2 (African) were associated with ischemic stroke. Circulatory diseases were overrepresented in discovery and replication cohorts with an early-onset of cardiovascular phenotypes irrespective of race. Sex- and race-dependent effect for cerebrovascular disease risk was clearly detectable, particularly for *NOTCH3(*HR_earlyonset_ = 2.175[1.391-3.403], p = 0.001), and *TREX*1(HR_earlyonset_ = 4.006[1.797-8.931], p = 0.001). *COL4A*1*/2*(HR_earlyonset_ = 2.163[0.87-5.38], p = 0.097). *HTRA*1 (European) also showed a similar trend of association.

**Conclusion:** Carriers for monogenic CSVD risk genes demonstrated the increased risk for the lifetime or early onset of cerebrovascular disease and diseases within or beyond the circulatory system, some of which in a race- and sex-dependent manner. Our findings support the concept of developing a gene panel of CSVD for population screening of patients with early-onset circulatory diseases.

**Key Points:** *Question:* Is there an increased risk for a lifetime or early onset of cardiovascular diseases and others within or beyond the circulatory system observed in rare variant carriers of seven well-characterized monogenic cerebral small vessel disease (CSVD) risk genes (*COL4A1, COL4A2, NOTCH3, HTRA1, TREX1, CTC*1, and *GLA*)?

*Findings:* In this retrospective longitudinal study of 175K individuals, which were temporally split into discovery (2738/5490) and replication cohorts (1695/3410) for carriers/matched noncarriers, *NOTCH3* (HR*_earlyonset_ = 2.175* for White), *TREX*1 (HR*_earlyonset_ = 4.006* for White), and COL4A1/2 (HR*_earlyonset_ = 2.163* for Black) were associated with ischemic stroke, particularly in female participants. *HTRA*1 also showed a similar trend. Circulatory diseases were overrepresented in discovery and replication cohorts with an early onset of cardiovascular phenotypes irrespective of race.

*Meaning:* Carriers for monogenic CSVD risk genes demonstrated the increased risk for the lifetime or early onset of cardiovascular disease and diseases within or beyond the circulatory system, some of which are race- and sex- dependent.

## 1. Introduction

Exome sequencing has made it possible to identify rare variants(RVs) in known genes causing monogenic stroke at both familial and population levels^1-4^. Previous attempts have been made to compile a comprehensive gene panel based on earlier linkage studies of monogenic stroke with Mendelian inheritance^5,6^. Whether used for familial^5^ or population^7,8^ screening, this technology, which combines exome capture and targeted sequencing, has demonstrated its potential to identify risk for monogenic stroke with early^5^ or late onset^7,8^.

Incomplete penetrance, variable expressivity, and pleiotropy lead to the phenotypic diversity^9^ and become potential obstacles to promoting exome sequencing in diagnosis of monogenic cerebral small vessel disease (CSVD). These obstacles include: 1) Limited knowledge of **lifetime incidence** of disease among individuals with RVs identified from risk genes; 2) **Subtype dependency** of risk genes leading to varied effect size^10^; 3) **Variable expressivity** of phenotypes in patients with rare heterozygous variants for monogenic diseases^11,12^; 4) **Incomplete penetrance** of pathogenic variants which could be modified by age for late-onset diseases^9^, even for autosomal dominant disorders^11,13-15^; 5) **Race- and sex-dependency** in monogenetic diseases with complex disease profiles^6,9,16^; 6) **Predisposition symptoms**^17^ (e.g. transient ischemic attack (TIA)^18^, hypercoagulability^19^, atrial fibrillation^20^), and patterns of comorbidities that have not been explored in individuals with RVs – leading to opportunities for personalized prevention strategy; 7) Development of a variety of small vessel-related phenotypes beyond the brain^21^ and circulatory system^22-24^ at the early stage in carriers (**Pleiotropy**). Finally, stroke is an ultimate expression of a systemic failure in arteriolar function^25^. Geisinger’s comprehensive Electronic Health Record (EHR) intersected with MyCode exome sequencing for genomic screening make it possible to identify potentially pathogenic variants in biobank-leveled population and associated disease profile and trajectory^1,11,26,27^.

The purpose of this study is to identify rare, potentially pathogenic variants of seven well-characterized monogenic CSVD risk genes (*COL4A1, COL4A2, NOTCH3, HTRA1, TREX1, CTC1*, and *GLA*) using a genotype-first approach within a healthcare system population-based biobank cohort. The goal is to explore the clinical manifestations observed in variant carriers and assess the risk for the lifetime or early onset of CSVD and other diseases within or beyond the circulatory system.

## 2. Method

### 2.1 Participating cohorts

This was a retrospective longitudinal study with a two-step design. The Geisinger patient population with comprehensive EHR was intersected with the MyCode initiative for exome sequencing of blood samples, which was performed through the DiscovEHR collaboration between Geisinger and the Regeneron Genetics Center (Tarrytown, New York) ^1,26,28,29^. The Geisinger Institutional Review Board approved this study to meet “Non-human subject research” using de-identified information. All research was performed in accordance with relevant guidelines/regulations. Informed consent was obtained from all participants and/or their legal guardian(s). The participants and analysis pipeline are illustrated in **Figure 1**.

**Figure 1.**
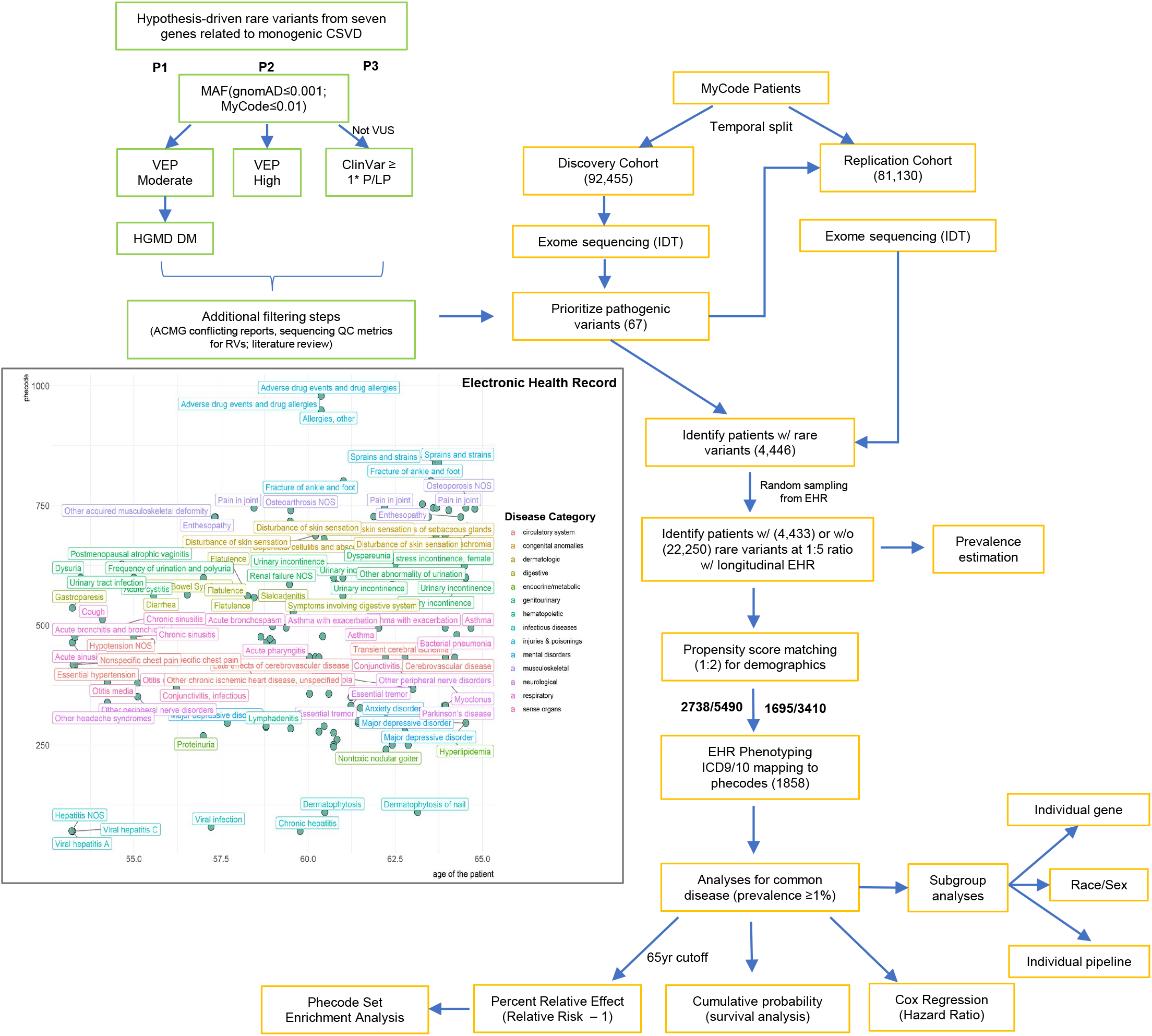
A flowchart of study design and analytic pipeline for the genotype-first phenome-wide longitudinal study. **A** diagnostic panel of RVs (n=67) prioritized by three pipelines (P1-3) was created for the purpose of interpreting exome sequencing results for monogenic stroke candidate genes (*COL4A1, COL4A2, NOTCH3, HTRA1, TREX1, CTC1*, and *GLA)*. Simulated longitudinal disease profile for a patient with RV(s) as an example to illustrate the data extraction and PheCode mapping. The colors represent different PheCodes from the same disease category. Cases for the specific PheCode were defined by ‘Rule of Two’ from any encounter types in the EHR.

The MyCode participants, a predominantly adult population with European ancestry, were temporally split into discovery (n=92,445) and replication (n=81,130) based on the time of dataset availability.

### 2.2 Genetic data

Probes (xGen Exome Research Panel v1.0.) from Integrated DNA Technologies (IDT) were used for the exome capture. Sequencing was performed by 75bp paired-end reads on Illumina HiSeq2500. Coverage depth was sufficient to provide more than 90% coverage for 99% of samples. Samples with low-quality sequence data or showing discordance of sex from self-reported sex or genotypes from array-based data were removed.

Alignments and variant calling were based on the human GRCh38 reference genome. Variant calls were produced by the WeCall variant caller (Genomics PLC, Version 1.1.2). A project variant-call file (pVCF) was compiled using the GLnexus^30^ joint genotyping tool (version 1.1.3-4). Sample level variant calls with site read depth (DP) <15, Phred-scaled genotype quality score (GQ) < 40, and alternate allele balance <15% (or fewer than 5 alternate reads) were removed. The call rate was 99.97%. Participants with the missing genotyping call (<0.001%) were considered null for the rare allele.

### 2.3 Prioritization of potentially damaging rare variants

RVs from seven risk genes for monogenic CSVD were identified from pVCFs by three variant annotation pipelines ^31^. They were *COL4A1* (OMIM:120130), *COL4A2* (OMIM:120090), *NOTCH3* (OMIM:600276), *HTRA1* (OMIM:602194), *GLA* (OMIM: 300644), *CTC1* (OMIM:613129), and *TREX1* (OMIM:606609) as shown in **Supplementary Figure 1**. Both *COL4A1* and *COL4A2* are located on 13q34. They are structurally expressed and typically function together. Thus, we treated them as one in this study. Variants were initially filtered by minor allele frequency (MAF) because the rate of pathogenic variation was lower due to selection constraints. Only variants with MAF < 0.001 from gnomAD and MAF < 0.01 from the MyCode population were considered as rare and subjected to the following variants annotation process. The MAF cutoffs were empirically determined based on the characteristics of each study population.

VEP annotation was conducted on 12/29/2019 using Ensemble web interface (https://useast.ensembl.org/info/docs/tools/vep/index.html), and the result in Supplementary Table 1 was updated on 01/24/2022.

Pipeline 1(P1): RVs with functional impact predicted to be ‘moderate’ by VEP annotation were subsequently filtered by HGMD (professional 2018.2 released) annotation^32^. Only variants predicted to be ‘disease modifying’ (DM) entered the final variant assessment process.

Pipeline 2(P2): RVs with functional impact predicted to be ‘high’ by VEP annotation were retained^33^. All of them are predicted loss-of-function (LoF) variants with enriched protein truncation variants (PTVs).

Pipeline 3(P3): RVs in ClinVar annotated as ‘pathogenic’ (P) or ‘likely pathogenic’ (LP), without conflicting assertions (as of 12/29/2019 database and updated on 01/24/2022 database) were selected^34^. ACMG interpretation criteria were applied^34^ and no VUS variants were included.

For variants with minor allele count (MAC) < 3 in the entire MyCode population, participant-level quality control (QC) data (GT, DP, AD, GQ, SBPV) from pVCF file or individual CRAM file for mapping/alignment were accessed by Integrative Genomics Viewer (https://software.broadinstitute.org/software/igv/home) to determine the final genotyping call.

Based on the above RV selection criteria(P1-3), we identified a total of 188 RVs in the discovery cohort. We further filtered out some RVs (n = 121) due to 1) conflict reports of significance based on ACMG criteria^34^; 2) more stringent sequencing QC score for rare frameshift variants; 3) clear evidence of benign variants reported by ACMG but identified by HGMD as ‘disease modifying’; 4) prior knowledge based on a literature search (*in vitro* functional studies as well as pedigree studies). Our preliminary study using *NOTCH3* as an example showed that adding this filtering process would help to enrich the pathogenic variants with higher penetrance (data not shown).

The prioritized 67 RVs, including their genotype counts as well as functional consequence (HGVSp) are illustrated in **Supplementary Figure 1** using UCSC Genome Browser (GRCh38/hg38). Additional tracks from ClinVar, HGMD, OMIM, and Pfam Domains (GENCODE database) were mapped to those variants.

### 2.4 Phenotype data

Geisinger billing codes for diagnostics at any encounter setting were mapped to the *International Classification of Disease (ICD)*-related ICD9 and ICD10 codes. Demographic information was extracted from the structured EHR. ICD-9 and ICD-10 diagnostic codes were combined and mapped to 1,866 hierarchical PheCodes^35^, each representing a specific disease phenotype. Rollup PheCodes without clinical definition of disease were removed from the subsequent analyses. All the PheCodes were grouped into 18 disease categories, such as the Circulatory system, the Genitourinary system, and more, as shown in a longitudinal layout (Figure 1). Study participants were labeled with a PheCode if they had one or more PheCode-specific ICD codes. “Case” was defined as all study participants with the PheCode of interest, and “control” was defined as all study participants without the PheCode of interest during their lifetime or within a predefined observation window for early-onset (e.g., ≤65yrs). To avoid an uneven sampling of the disease trajectory, which might lead to temporal bias in the longitudinal study, we also tested the age cutoff at 60 years or 55 years. Sex, as a biological variable, was included during the mapping process so that sex-specific PheCodes would not be mislabeled.

### 2.5 Comparability of variant carriers and non-carriers

Propensity score matching (PSM) to control confounding was applied to obtain the ‘counterfactual’ control group. Race, sex, and index age, which were denoted as the last active (last encounter) date, were identified as potential confounding factors. Other unmeasured confounding factors were assumed to be correlated with these factors and their confounding effects would be largely reduced after matching. They were selected as covariates in a Logistic Regression model to create propensity scores (R ‘MatchIt’ package). We chose “nearest neighbor matching without replacement” to create a more balanced carrier:noncarrier ratio at 1:2, as shown in **Supplementary Figure 2**; this step ensured that the standardized mean differences (SMD) for the matched variables were within ± 0.1. The number of participants in both cohorts was 2738/5490 and 1695/3410, respectively. For noncardiovascular-related common diseases, Risk Differences (RD) were < 0.1 for lifetime and early onset (≤65yr) in both cohorts, suggesting no selection bias between the carrier and noncarrier groups^36^.

### 2.6 Outcome measures

Major outcome variables in this study included age of onset, relative risk (RR) and percent relative effect (PRE).^37^

Risk Ratio (Relative Risk, RR) is defined as, 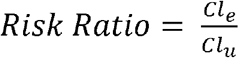, where (*Cl*_*e*_) = cumulative incidence among the exposed subjects, and (*Cl*_*u*_) is the cumulative incidence among unexposed subjects. The relative risk (or risk ratio, *RR*) is an intuitive way to compare the risks for the two groups.

*RR* -1 is defined as the percent change in the exposed group (carriers). We consider the unexposed group (noncarriers) as having 100% of the risk, and the phenotype expression of the exposed group, expressivity, is relative to that. *PRE*, or *RR* – 1, is alternatively interpreted as a normalized *Rn* with a positive or negative value. The *PRE* for each phenotype after exposure is similar to the percentage change in gene expression after exposure. This parameter will be used for PheCode set enrichment analysis, a counterpart of gene set enrichment analysis, to calculate the enrichment score.

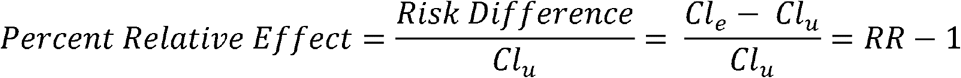

A circo plot was created based on the top 50 PheCodes ranked by *PRE* and clustered by the processes. This representation shows the extracerebral disease manifestations and overrepresentation while the rainbow links represent the association between PheCodes in carriers.

### 2.7 Statistics

Categorical data are reported as number (percentage); continuous data are presented as mean (SD), as indicated. Statistical analyses were performed using R, version 3.6. All mapped PheCodes were divided into common (with a better mapping performance^38^) or rare diseases by the prevalence (<1%) of the disease estimated from noncarriers. Fisher’s exact test was conducted to determine the frequency difference between PheCodes in carriers and noncarriers. Due to imbalanced case-control ratios for rare diseases which may lead to inflated statistical significance, only the associations between common diseases were reported. There were 756 common PheCodes mapped for 4433 participants, in which 695 or 580 phenotypes had case-control ratios greater than 1:100 (regarded as common diseases) for both cohorts, respectively.

Survival analyses and Kaplan-Meier estimator for cumulative incidence probability were conducted (R ‘survival’ and ‘survminer’ packages). Participants were considered at risk of diseases at lifetime (for up to 85 years of follow-up) or early onset (age of onset > 65 years was considered as null and were censored at the last active date or the date of data extraction (06/12/2021). Multivariate Cox proportional hazards model, adjusted for confounding variables (sex, index age, and 5 main principal components from PCA) was also performed and the hazard ratio for phenotypes of interest at lifetime as well as early-onset (≤65 years) was calculated. The proportional hazards assumption was verified using Schoenfield residuals. For all analyses, p<0.05 was considered statistically significant. For all post-hoc pairwise tests, p-values were present as raw.

### 2.8 Subgroup analyses

Subgroup analyses were conducted by stratifying the participants based on race (EUR or AFR), sex, and gene. The early onset was defined as patients with the first diagnosis before 65 years old. We also tested 60 and 55 years as cutoffs to determine if the trend of association or expressivity (*PRE*) remained valid. We evaluated the effect size (Hazard Ratio, *HR*) of RVs associated with PheCodes or PheCode sets in a multivariate CoxPH model in all samples as well as subgroups stratified by sex, race, and gene. All subgroup analyses were considered exploratory and p-values were present as raw.

### 2.9 PheCode Set Enrichment Analysis (PSEA)

PheCode Set Enrichment Analysis was conducted to determine which PheCode set from a disease category (e.g., circulatory system) was significantly enriched against a randomly selected PheCode set (same number) from that category. In this study, we only focused on common diseases, the prevalence of which in the MyCode population ≥ 1%. Using pre-ranked differential gene expression data to conduct gene-set enrichment analysis (GSEA) inspired us to develop the PSEA. The general idea of the PSEA method is to test whether the distribution of PheCodes in the PheCode-set differed from a uniform distribution. To achieve this goal, we used the established ranking metric strategy used in GSEA – namely, signed statistic (e.g., disease-level *PRE*) was selected as a substitute to the statistics of differential gene expression used in GSEA.

fgsea^39^ is an R-package for fast pre-ranked gene set enrichment analysis. (https://github.com/ctlab/fgsea). Here we adopt the idea and procedure of GSEA. The preranked PheCode-level statistics S (PRE) for the PheCode U = {1, 2, …, N} and a list of query PheCode sets, P, resulting in Max(ES) per disease category. This analysis aims to determine which of the PheCode sets from P has a non-random behavior. Fgsea used a gene set enrichment score function *S*_*r*_ (*p*) originally defined by Subramanian et al^40^. The more positive is the value of *S*_*r*_ (*p*) the more enriched the gene set is in the positively-regulated genes (with *S*_*i*_ > 0). Accordingly, negative *S*_*r*_ (*p*) corresponds to enrichment in the negatively regulated genes. Value of *S*_*r*_ (*p*) can be calculated as follows. Let *k* = |*p*|, *NS* = ∑_*i*ϵ*p*_ |*S*_*i*_|. Let also ES be an array specified by the following formula^39^:

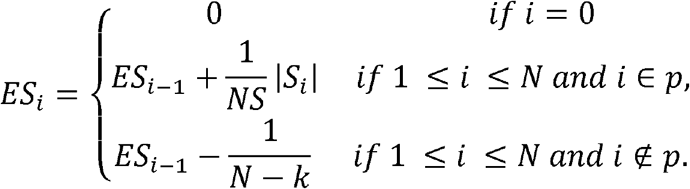

The value of, *S*_*r*_ (*p*) correspond to the largest by the absolute value entry of ES^39^:

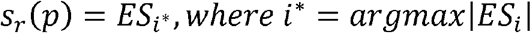

Following Subramanian et al. ^40^ for a pathway, p, Korotkevich et al. define GSEA p-value as^39^:

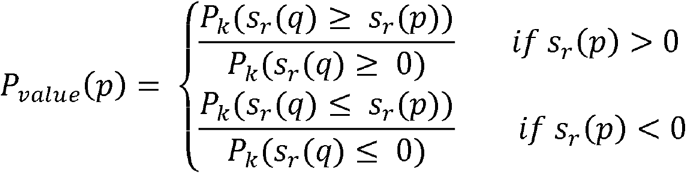

Where *q* is a random gene set of size *k*.

NES = enrichment score normalized to mean enrichment of random samples of the same size of tested PheCode-set. P-value estimation is based on an adaptive multi-level split Monte-Carlo scheme, which allows for estimating a lower p-value (< 10^−5^). To speed up the process, fgsea divides the entire ranked PheCodes into n blocks and tries all the thresholds to get the minimum p-value. P-value adjustment for multiple testing of a number of PheCode sets was conducted using the Benjamin-Hochberg procedure.

The codes are available at GitHub - TheDecodeLab/Trajectory-of-Monogenic-Diseases: Development of a platform to observe and analyses the trajectory of monogenic diseases.

## Result

### Participants Demographics

The entire dataset was temporally split based on the availability of the sequencing data. Demographic and clinical features for the discovery and replication cohorts are listed in **Table 1a**. We identified a total of 4,446 RV carriers of at least one of the seven candidate genes out of 173,575 MyCode participants. Initial EHR data extraction was completed on 12/29/2019 for a pilot study of the discovery cohort; the second EHR data extraction was completed on 12/06/2021 for both discovery and replication cohorts.

**Table 1.**
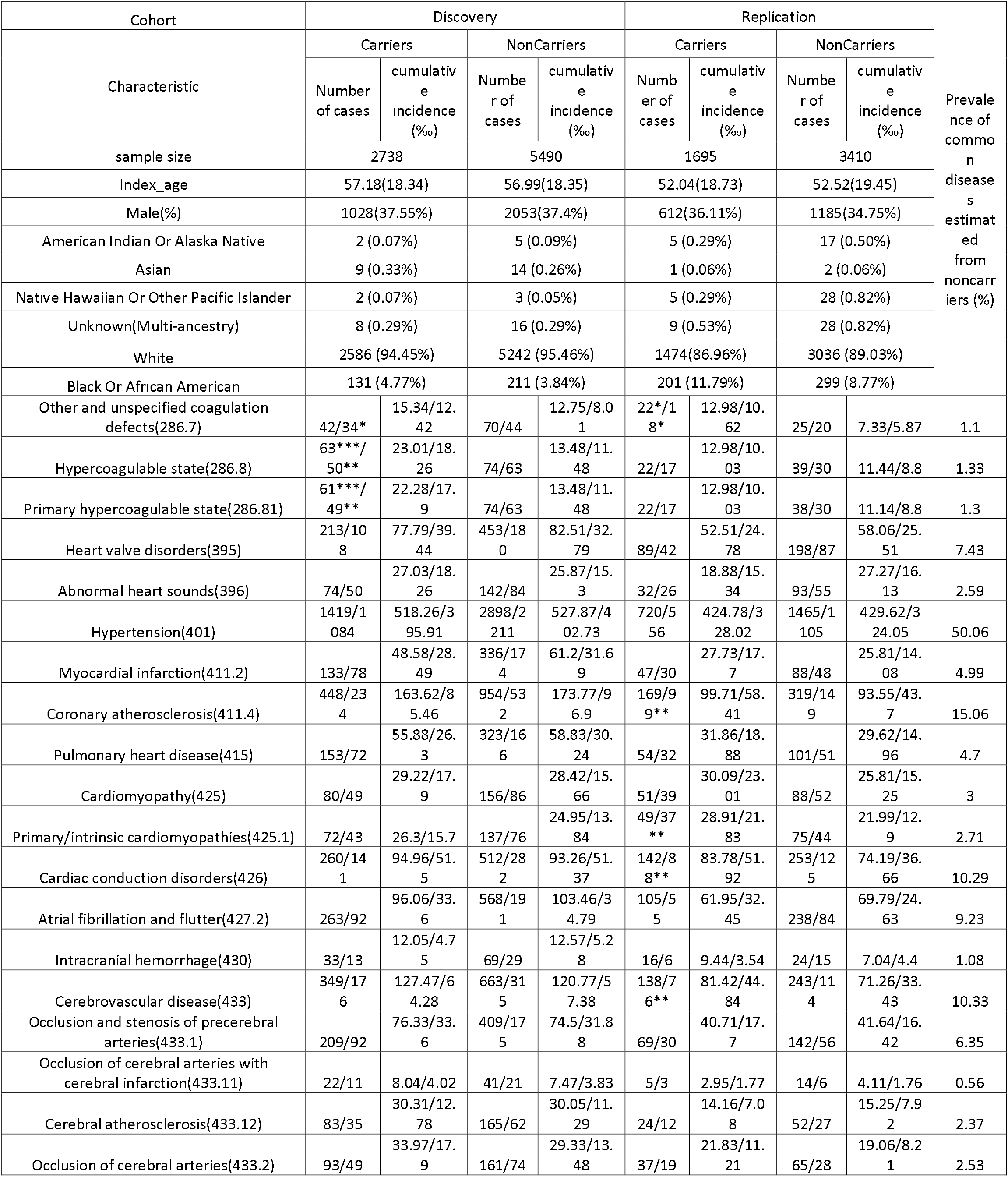
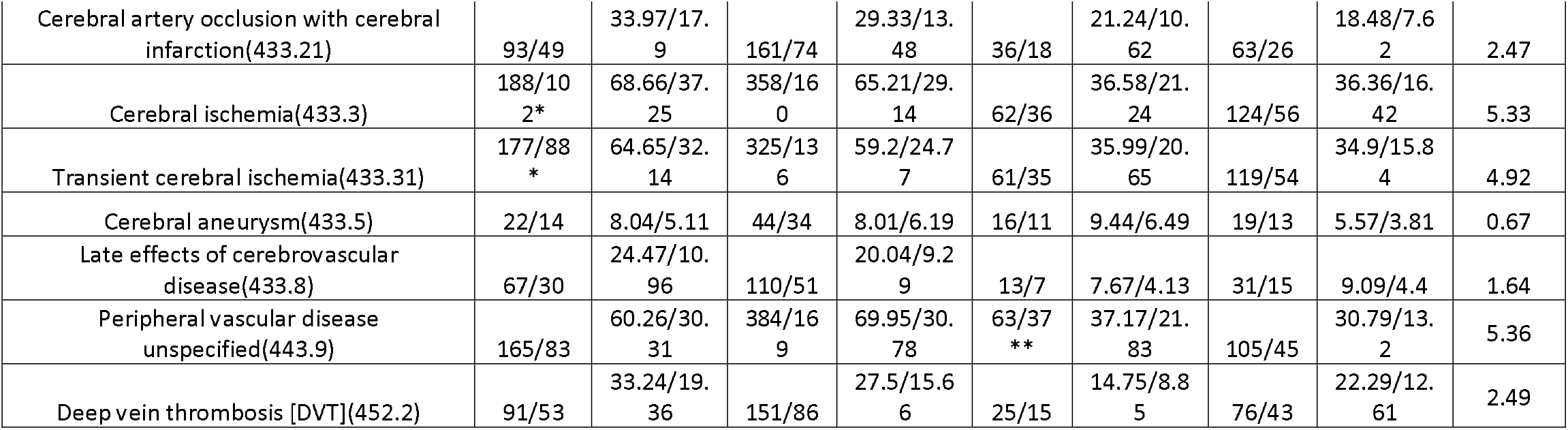
Patient characteristics of RVs carriers or noncarriers in both Discovery and Replication cohorts. The study cohort was split into Discovery and Replication datasets and further split according to having RVs or not. All continuous variables were reported as mean (standard deviation), and dichotomous variables were reported as absolute count (frequency). For disease phenotype, we reported the absolute count of patients having lifetime and early-onset of the corresponding PheCode from Geisinger EHR. For the lifetime observation, patients were followed up to 85 years; Patients with age of onset after 65yrs considered null for the early-onset cases. ANOVA or Fisher’s exact test was selected to determine the significant difference between groups for quantitative or qualitative measures, respectively.

After removing carriers without longitudinal EHR data (at least two encounters separated by at least one month), we end up with 4433 (2738/1695) carriers. Five-fold noncarriers, 22,250 (13,725/8525) were randomly sampled from noncarriers in each cohort. The prevalence of any diseases in the MyCode population was estimated by these randomly selected noncarriers. After PSM at a 1:2 ratio, there was no significant difference in sex, race (EUR or AFR), or the index age between carriers and noncarriers. The replication cohort was, on average, five years younger than the discovery cohort. This difference may lead to some findings from the discovery cohort not being replicated in the younger cohort, particularly for late-onset diseases.

### Distribution of RVs identified by three pipelines

A total of 24 participants had RVs from any two genes. Around 20.18% (569/2738 and 329/1695) or 40.61% (1244/2738 and 563/1695) of participants had at least second/third-degree relatedness based on PiHAT score of 0.20 or 0.10, respectively. Most participants (94.45%/86.96%) are self-claimed White and confirmed with EUR ancestry by Principal Component Analysis (PCA). A smaller portion of participants (4.77%/11.79%) are self-claimed Black and confirmed with AFR ancestry by PCA. Participants from other ethnical groups were < 2% and were not included for further analysis.

Missense (38 distinct variants), stop gain (12), NMD (nonsense-mediated decay), transcript variant (8), and splice donor (5)/acceptor (7) variants were the top categories of VEP annotated consequence (**Supplementary Table 1**), suggesting these potential pathogenic variants may have an impact on protein function at the various levels.

In **Table 1a**, most participants with RVs were female in both cohorts. RVs were mainly identified in *COL4A1/2* and *NOTCH3* genes (**Table 1b**).

According to PSM plots (**Supplementary Figure 2**), RVs were observed disproportionately more in participants with AFR ancestry (for the unadjusted sample, SMD deviated to the right side for AFR, whereas SMD deviated to the left side for EUR) in both cohorts. The majority of identified RVs were in heterozygous form. Compared to the replication cohort, the discovery cohort had more participants (EUR: 60.70% vs. 38.54%; AFR: 95.42% vs. 89.05%) with *COL4A1/2* variants but fewer participants (EUR:22.37% vs. 34.57%; AFR: 0% vs. 2.49%) with *NOTCH3* variants. The distributions of RVs from the other four genes across the two cohorts were comparable (p > 0.05 by Fisher’s exact test) (**Table 1b**).

Three variant annotation pipelines were applied to capture potentially damaging RVs. RVs derived from P2 were enriched with putative loss of function variants affecting RNA splicing (splice donor and acceptor variants) or translation (start lost, stop gained, and stop lost). However, the replication cohort had fewer participants identified from pipeline 2 with these consequences (**Supplementary Figure 3**). It was noted that many of the RVs prioritized by P2 were singleton or doubleton with acceptable QC data. Using PheCodes related to the circulatory system as examples, we explored the performance of the three pipelines. These PheCodes showed a stronger percent relative effect (*PRE*) in either discovery or replication cohorts (**Supplementary Figure 3**). P1 showed the largest *PRE* (green bar) in most of the PheCodes than other pipelines or the intersect pipeline (red bar), suggesting that P1 augmented the performance of the P3 (purple bar), the ClinVar annotation. A similar observation was reported by others^41^. In some PheCodes, P2 (blue bar) showed the largest *PRE*_*S*_, particularly in the discovery cohort but less obvious in the replication cohort, perhaps due to fewer carriers (44% versus 10%) exclusively identified by P2. Since three pipelines show their capability of identifying positive cases (*PRE* ≥ 0) for some but not all tested PheCodes at varying degrees, carriers identified from all pipelines were combined for the following study.

### Top PheCodes with the largest *PRE* were overrepresented in early-onset patients

The circo plot (**Supplementary Figure 4**) demonstrated the top 50 PheCodes ranked by *PRE*, grouped by the process. Unlike AFR participants with overrepresented phenotypes in the circulatory system, EUR participants did not show any disease categories represented excessively in the top 50 PheCodes with increased lifetime risk. Only in early-onset patients, diseases from the circulatory system were exclusively overrepresented in the top 50 PheCodes in both EUR and AFR participants (**Figure 2**). Primary cardiomyopathy was the only shared PheCode in the circulatory system in early-onset EUR between discovery and replication cohorts, whereas several PheCodes related to cerebral vascular diseases were top-ranked in the lifetime and early-onset AFR cohorts.

**Figure 2.**
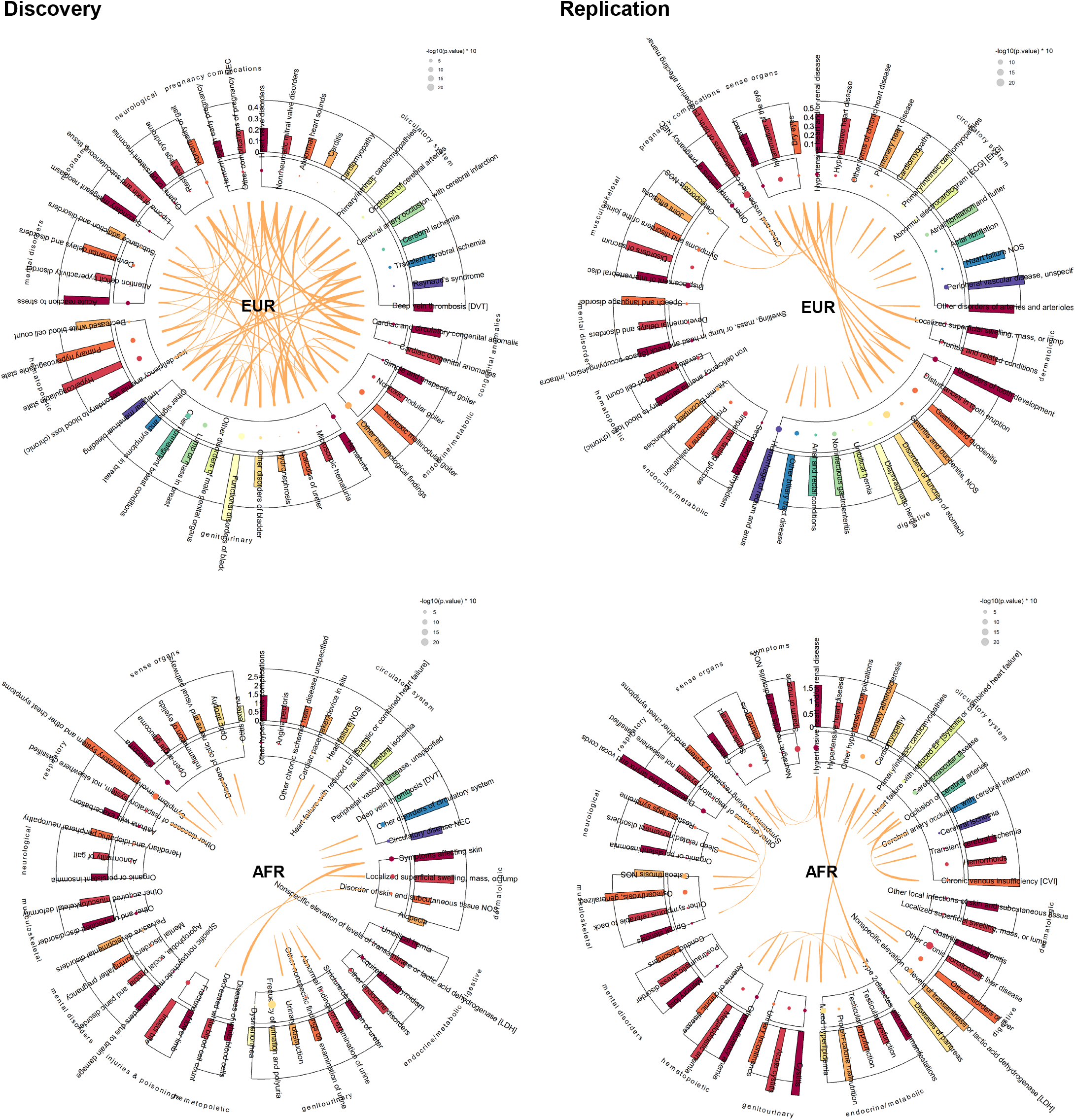
Circo plots with interlinks of the top 50 common diseases of which RVs from seven CSVD risk genes having increased relative risk in early-onset patients. Participants with the age of onset for a specific disease after 65yrs were considered null for the early-onset phenotype; A circo plot was created based on the top 50 PheCodes ranked by precent relative effect (PRE) and clustered by the processes. The links represent the significant association between two PheCodes in RV carriers calculated by Fisher’s exact test after correction for multiple testing (p < 0.001 given 50 phecodes being tested)). Significant associations for adjacent PheCodes were labeled with spike but not linked. Y axis represents PRE; The area of the dot was proportional to the significant association between carriers and the corresponding disease according to Fisher’s exact test. Diseases from the circulatory system were overrepresented in early-onset carriers in both cohorts with EUR or AFR ancestry. Disease categories with the number of PheCodes >= 2 were plotted.

It was noted that all PheCode links between and within categories were specific to carriers in both AFR cohorts since no links were identified in noncarriers. However, some links identified in both EUR cohorts were shared by carriers and noncarriers. For example, the links between cardiac and circulatory congenital anomalies and heart valve disease were identified in both carriers (Figure 3), particularly for *COL4A1/2* **(Supplementary Figure 5)** and noncarriers.

**Figure 3.**
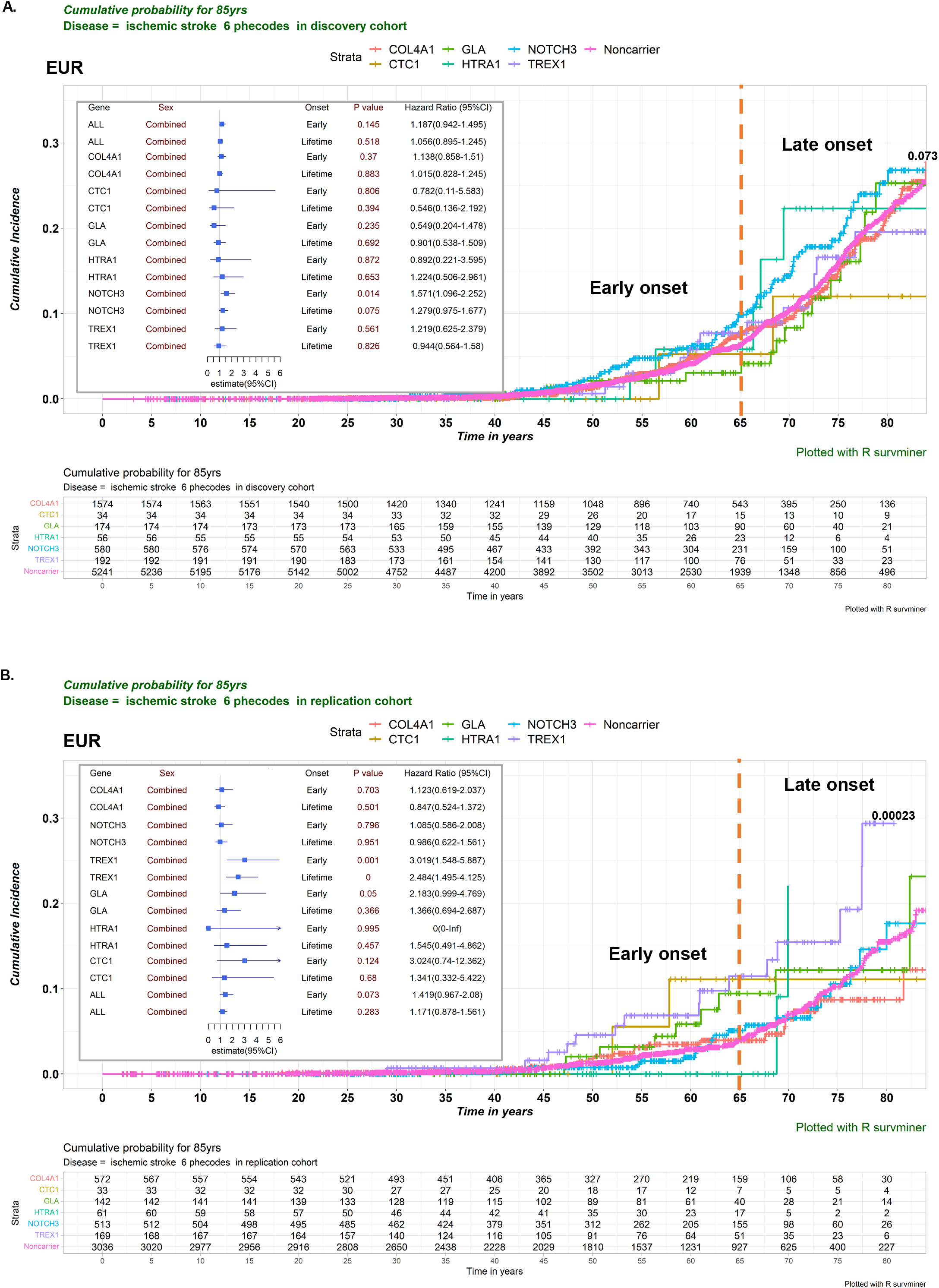
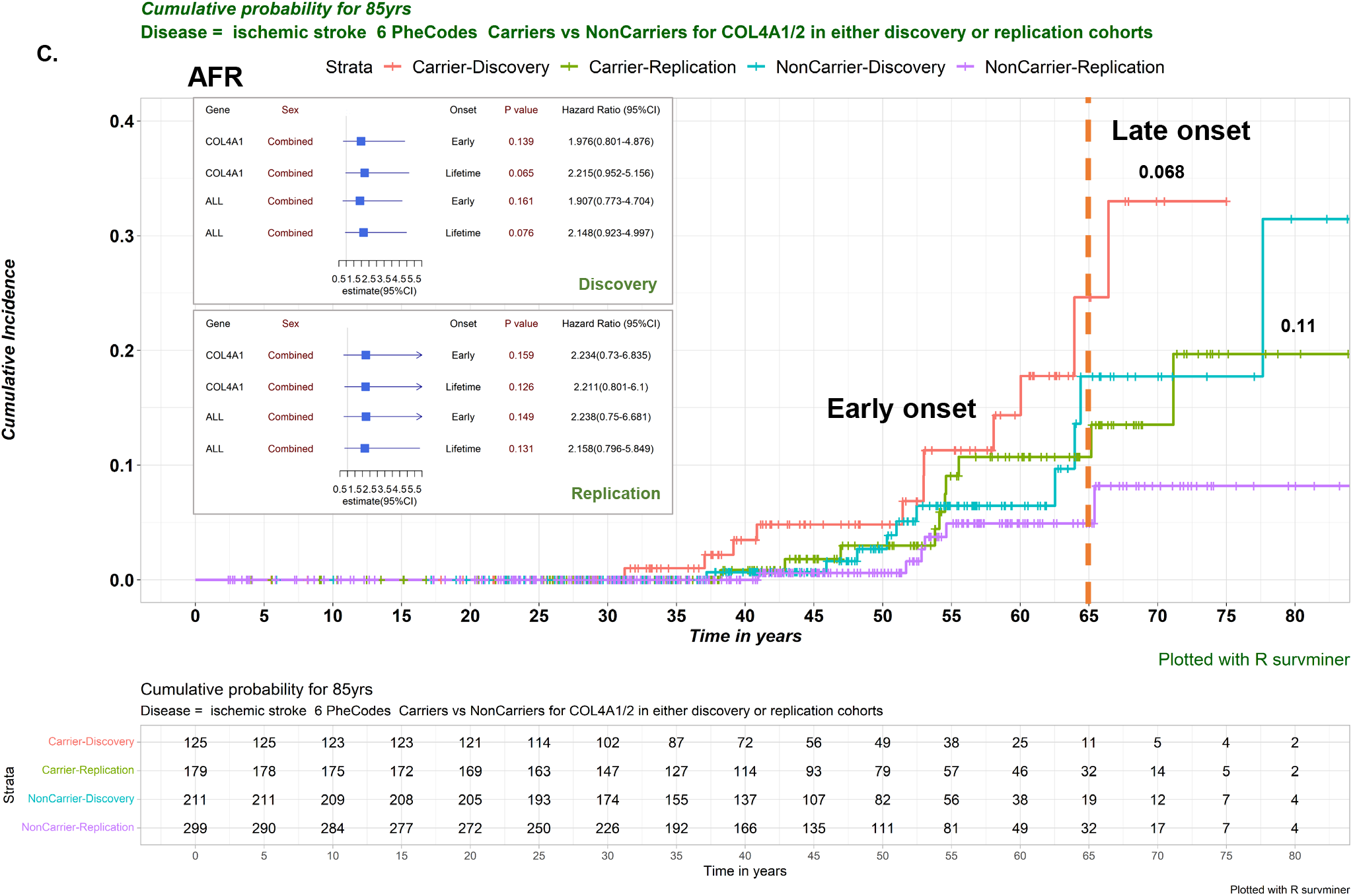
Kaplan-Meier estimated the cumulative probability of ischemic cerebrovascular disease for RV carriers compared to matched noncarriers, a gene- and race-dependent association. **A**. EUR in discovery cohort; **B**. EUR in replication cohort; **C**. AFR in discovery/replication cohort. Carriers for genes other than COL4A1/2 were too low to show in AFR subgroups. Only patients with European (EUR) or African (AFR) ancestry were included in this study. Ischemic cerebrovascular disease was determined by six PheCodes. The follow-up was censored on 12/06/2021. A Lifelong (up to 85yrs) Log-rank test was applied to comparing carriers for each gene to the shared noncarriers for all seven genes. For post-hoc pairwise tests, p-values were present as raw. Cox proportional-hazards regression model, adjusted for sex, index age, and five principal components, was conducted to determine the association between collapsed RVs from each gene or combined and the IS. Adjusted Hazard Ratios (HR) estimates and their 95% confidence intervals (Cis) for a lifetime (up to 85yrs) or early-onset (≤65yrs) risk were present in the forest plot.

For individual genes (e.g., *COL4A1/2* and *NOTCH3*) in the early-onset study, PheCodes from the circulatory system were overrepresented in the top 50 common diseases in both discovery and replication cohorts (**Supplementary Figure 5**). However, this observation was valid only in *TREX1 and CTC1* in the replication cohort. The increased risk for cardiac and circulatory congenital anomalies^22,23 42^ was identified in the EUR discovery cohort (**Figure 2**), particularly for *COL4A1/2* **(Supplementary Figure 5)**.

Some connections between CSVD genes with phenotypes beyond cardiovascular systems were captured. These secondary findings included: 1) *GLA* and thyroid diseases; It was previously suggested that a physiological concentration of thyroid hormone directly facilitates matrix Gla protein gene expression in smooth muscle cells via thyroid hormone nuclear receptors, leading to prevention of vascular calcification ^24^; *COL4A1/2 and HTRA1* with renal findings (hematuria)^23^, and *COL4A1/2* with abnormal function of bladder^43^; 3) *HTRA1* and musculoskeletal disease^44^.

Notably, some common cerebrovascular disease-related phenotypes (such as TIA, primary cardiomyopathy, atrial fibrillation, and hypercoagulability) were top ranked with an increased relative risk in at least one cohort. The links between TIA and cardiomyopathy and cerebral ischemia and carditis were captured in the discovery cohort of EUR carriers but not in noncarriers. The link between occlusion of cerebral arteries and chronic venous insufficiency was identified only in AFR carriers but not in noncarriers in the replication cohort.

### Race-, and Sex-dependent association

The entire gene-specific survival analysis is illustrated in **Figure 3**. In the discovery cohort, *COL4A1/2* or *NOTCH3* variants showed the (trend of) association with IS in AFR or EUR participants, respectively. In the replication cohort, *COL4A1/2 or TREX1* variants showed a similar (trend of) association with IS in AFR or EUR participants, respectively. When combined both AFR cohorts (**Figure 4C**), *COL4A1/2* was significantly associated with IS (HR_lifetime_=2.032(1.065-3.875), p=0.031). It was noted that *HTRA1* carriers had an increased cumulated incidence of IS in the late-onset participants from both discovery and replication cohorts (the green curve spikes after 65yr in **Figure 3A-3B**).

**Figure 4.**
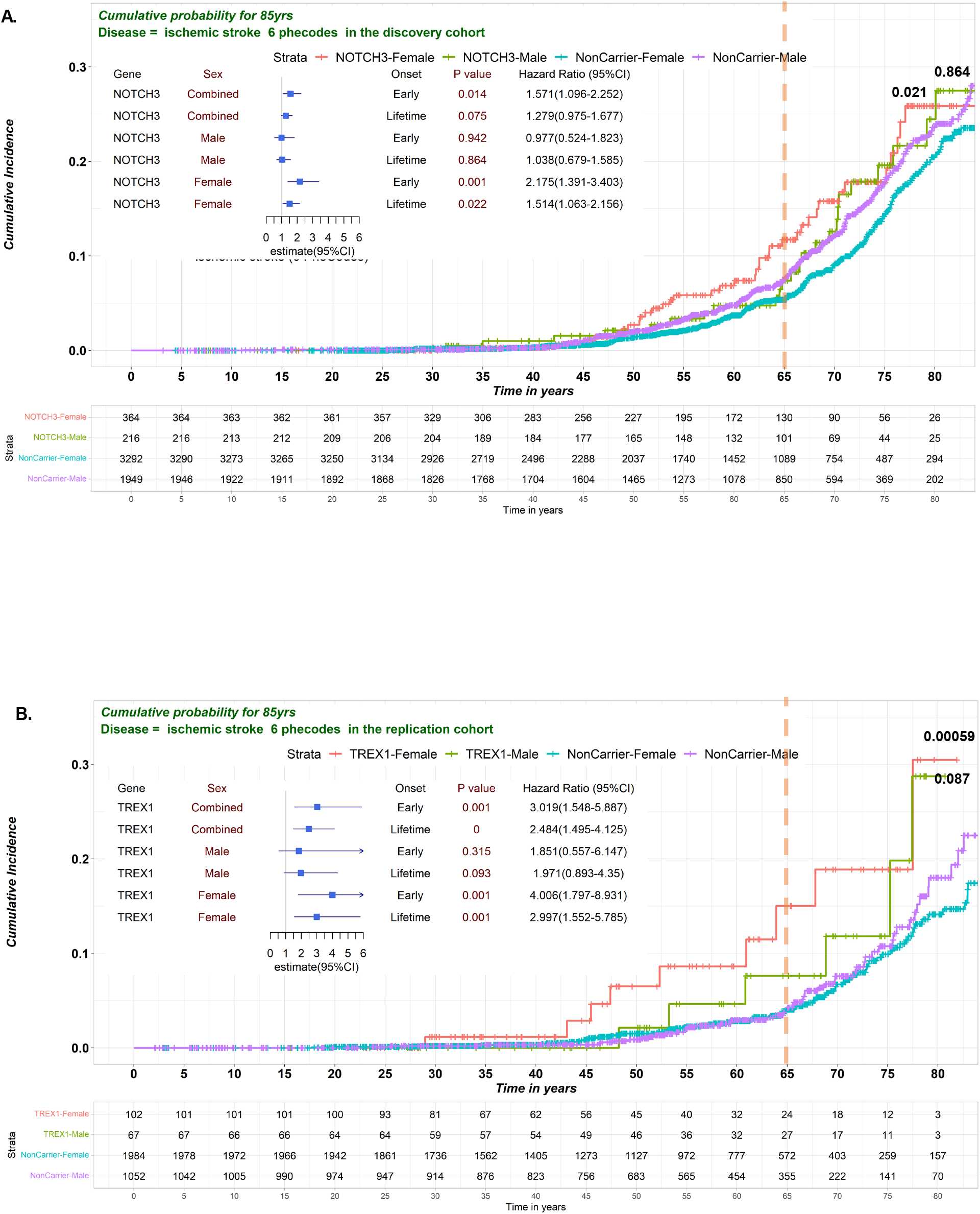
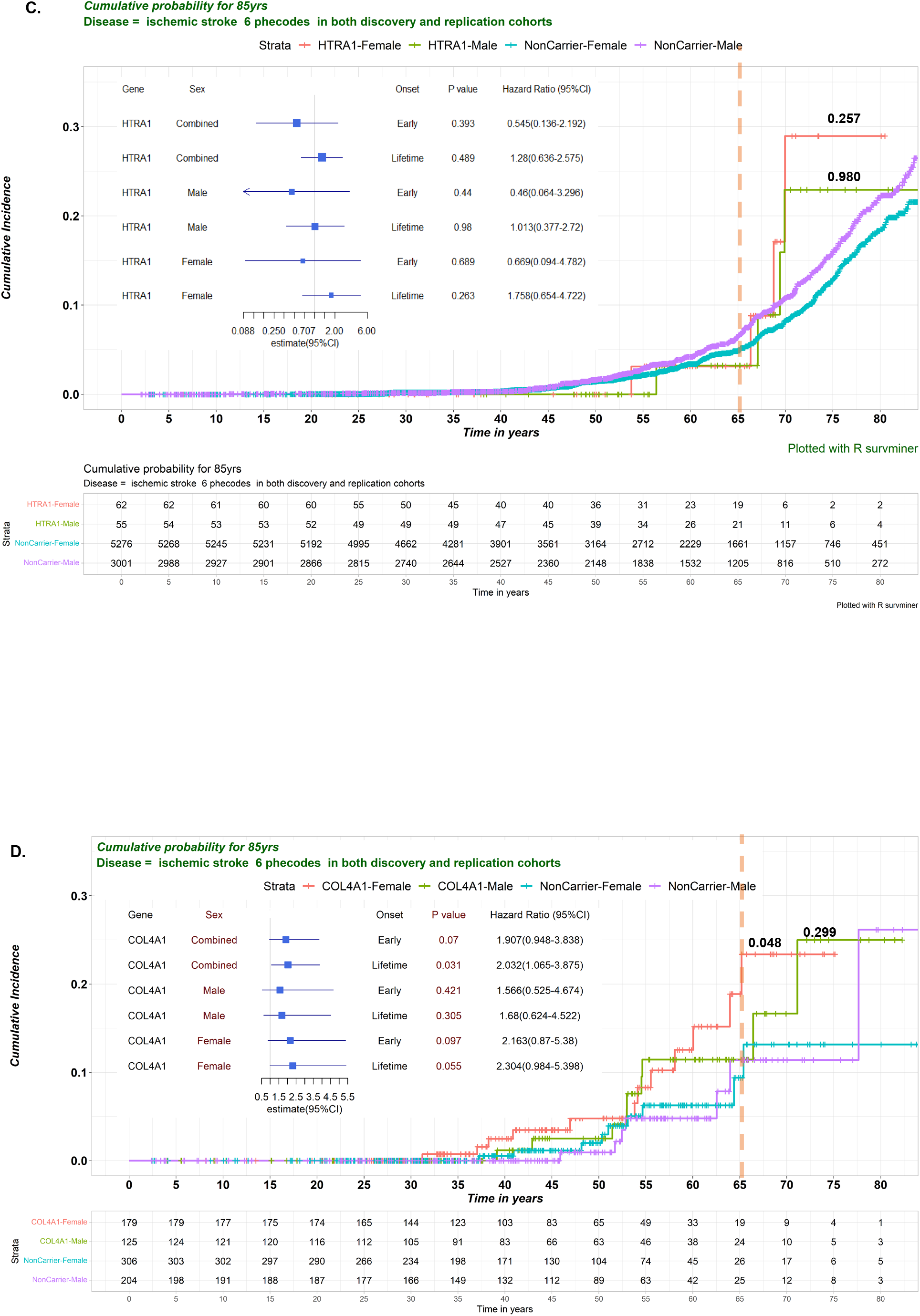
Kaplan-Meier estimated the cumulative probability of IS for carriers compared to matched noncarriers, a sex-dependent analysis. For post-hoc pairwise Log-rank tests, p-values were present as raw. The effect size (HR) was observed to be larger in Female than Male carriers. A. NOTCH3 in the discovery EUR cohort; B. TREX1 in the replication EUR cohort; C. COL4A1 in both discovery and replication AFR cohort. Cox proportional-hazards regression model, adjusted for sex, index age, and five principal components, was conducted to determine the association between collapsed RVs from each gene and the IS. Adjusted Hazard Ratios (HR) estimates and their 95% confidence intervals (Cis) for a lifetime (up to 85yrs) or early-onset (≤65yrs) risk were present in the forest plot.

Sex-dependent association was demonstrated in **Figure 4**. In the discovery cohort, the effect sizes were larger in female participants (HR_earlyonset_=1.499[1.046-2.148], p=0.027 for *COL4A1/2* and 2.175[1.39-3.401], p < 0.001 for *NOTCH3* in EUR; 2.343[0.713-7.701], p=0.161 for *COL4A1* in AFR). In the replication cohort, female patients with early-onset IS showed larger effect sizes (HR_earlyonset_=2.445[0.584-10.232], p=0.221 for *COL4A1/2* in AFR; 4.006[1.797-8.931], p<0.001 for *TREX1* in EUR). However, these findings for *NOTCH3* or *TREX1* were not observed in the replication or discovery cohort, respectively. AFR participants had no RVs identified for *NOTCH3*. This sex-dependent effect also appeared in *HTRA1* carriers after combining samples from both discovery and replication cohorts with a trend of increased effect size in female carriers (HR_lifetime_=1.758[0.654-4.722], p=0.263) but not in male carriers HR_lifetime_=0.98[0.377-2.72], p=0.98).

*COL4A1/2* variants were associated with hypercoagulability in EUR participants in the discovery cohort (HR_lifetime_=1.49[1.065-2.084], p=0.020). The trend of association was observed in the replication cohort (HR_lifetime_=1.626[0.856-3.085], p=0.137) (**Supplementary Figure 7A**). Effect sizes were larger for *COL4A1/2* (HR_earlyonset_=1.963[1.285-2.998], p = 0.002) in EUR female participants of hypercoagulability, but not for *NOTCH3* (HR_earlyonset_=1.194[0.57-2.501], p = 0.638) in the discovery cohort. However, this significant association was only observed in the replication male cohort (HR_earlyonset_= 5.113[1.649-15.854], p = 0.005). The association between *COL4A1/2* variants and hypercoagulability was also identified in male AFR participants (HR_earlyonset_=5.113[1.649-15.854], p = 0.005). No connection between hypercoagulability and cerebrovascular disease was observed by Fisher’s exact test in all genes (**Figure 2**) or *COL4A1/2*-only (**Supplementary Figure 5**) in the discovery cohort. The connection between hypercoagulability and heart valve disorders was only observed in carriers (**Figure 2 and Supplementary Figure 5**) but not in noncarriers (data not shown) in the EUR discovery cohort. The association between HTRA1 and hypercoagulability was identified in the discovery EUR cohort and validated in the replication EUR cohort (**Supplementary Figure 7**).

*CTC1* had a significant association with primary cardiomyopathies[425.1] with a stronger signal in the replication EUR cohort (HR_earlyonset_=8.463[2.608-27.466], p=0.0006; HR_lifetime_=4.601[1.433-14.674], p=0.010) than in the discovery EUR cohort (HR_earlyonset_=4.568[1.118-18.663], p=0.034; HR_lifetime_=2.779[0.883-8.752], p=0.081) (**Supplementary Figure 7B)**. *TREX1* only demonstrated a significant association with primary cardiomyopathies in the replication EUR cohort (HR_earlyonset_=3.146[1.403-7.054], p = 0.005; HR_lifetime_=2.386[1.184-4.810], p=0.015).

### PheCode-set Enrichment Analysis in early-onset patients

The top enriched disease category was circulatory system in both discovery (N = 18, NES = 2.95, p_adj_ = 2.8×10^−10^ for EUR; N = 20, NES = 2.43, p_adj_ = 2.7×10^−9^ for AFR) and replication cohorts (N = 20, NES = 2.22, p_adj_ = 2.8×10^−7^ for EUR; N = 20, NES=2.36, p_adj_ = 8.5×10^−8^ for AFR (**Figure 5 and Supplementary Figure 8**). Validation across cohorts or across racial groups was also performed using the PheCode set derived from circulatory system as an example (**Figure 5D**). Neither the replication EUR cohort (NES = 1.10, p_adj_ = 0.97) nor the discovery AFR cohort (NES = 1.61, p_adj_ = 0.12) validated the enriched PheCode set from the discovery EUR cohort. The enriched PheCode set from the discovery AFR cohort was marginally replicated in the replication AFR cohort (NES = 1.72, p_adj_ = 0.083), but was not replicated in the discovery EUR cohort (NES = 0.77, p_adj_ = 0.96). The enriched PheCode set from the replication EUR or AFR cohort was mutually replicated each other (NES = 2.02, p_adj_ = 7.1×10^−4^ or NES = 1.89, p_adj_ = 1.6×10^−3^ in the replication AFR or EUR cohort, respectively).

**Figure 5.**
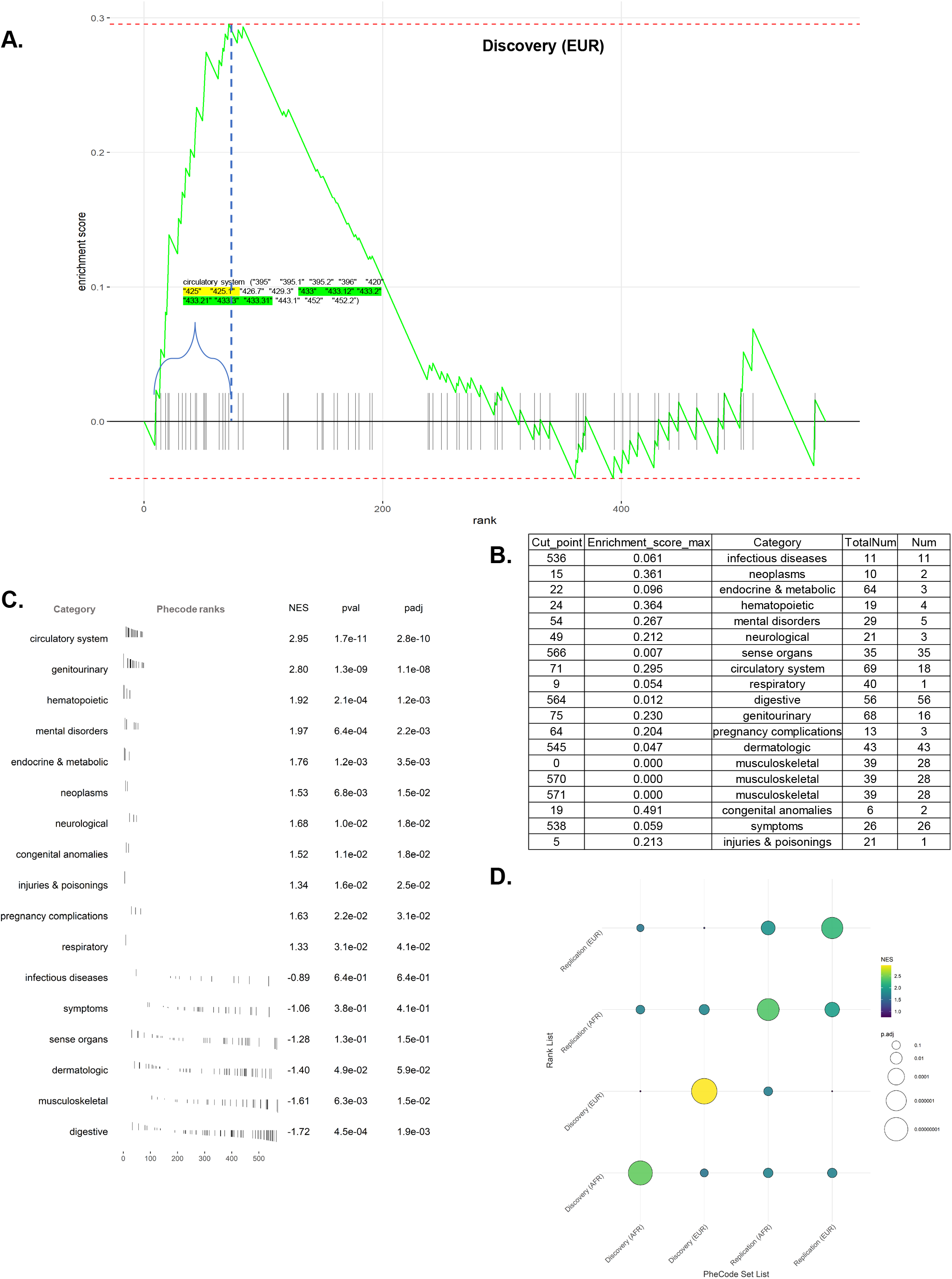
PheCode Set Enrichment Analysis was conducted to determine which disease categories were significantly enriched. **A**. A graph that corresponds to a calculation of enrichment score. Each breakpoint on a graph corresponds to a PheCode present in the disease category (e.g., Circulatory system). The curve goes rightward when phecode. disease category and goes upward when ∈ phecode sets. **B**. For example, in Discovery(EUR), the point corresponds to PheCode ranked the 71th from the entire PheCode of common diseases of any category, where the maximal value of the enrichment score is reached. The vertical bars along the x-axis represent 69 PheCodes belonging to the interrogated disease category(e.g., the circulatory system). The vertical dash line defined “leading edge subset” as all those PheCodes (in bracket) ranked as least as high as the enriched set, and the enriched score reached the maximum. **C**. The forest plot was created to show the effect size of significantly enriched PheCode sets from each disease category in Discovery(EUR), where the common diseases were ranked based on Percent Relative Effect (PRE). The raw and adjusted p-value for multiple testing using BH procedure was listed. **D**. Validation across cohorts or across races using the PheCode set derived from the circulatory system as examples. The PheCode set enrichment analysis has two input objects: 1) an array of pre-ranked PheCode-level statistics *S* (*PRE*) for the PheCode *U* = {1, *2*,, *N*}, shown as Rank List (y-axis); 2) a list of query PheCode sets *P* which consist of 18 enriched PheCode sets from the corresponding 18 disease categories. The goal of the analysis is to determine which of the 18 PheCode sets enriched from one dataset also shows a non-random overrepresentation in the other dataset. Here we only reported the result of PheCode Sets derived from the Circulatory system (x-axis).

## Discussion

There is an unmet need to evaluate the lifetime risk and the variability in age of onset for cerebrovascular disease by leveraging large-scale biobank datasets linked to comprehensive EHRs, and to identify potentially actionable genomic variants that may contribute to monogenic IS. As the management of individuals with monogenic causes for their diseases could be different from patients with polygenic or non-genetic etiology, this diagnostic attribution in common disease could alter the care options^17^.

In this study, using a workflow to prioritize potentially pathogenic variants by an integration of three variant annotation pipelines, we showed that 1) Most of the RVs identified were in a heterozygous form and disproportionately present in AFR participants; 2) Inclusion of participants with ultra-RVs, which predicted to have a ‘high’ functional impact and have not been observed in ClinVar and/or HGMD, may augment the disease profiling; 3) *NOTCH3, TREX1, and COL4A1/2* were associated with IS in a race- and sex-dependent manner (more frequent among women); 4) Circulatory diseases were overrepresented in discovery and replication cohorts with early-onset of cardiovascular phenotypes irrespective of race; 5) Carriers of the seven genes showed increased risk for early signs and symptoms of cardiovascular disease such as, TIA, cardiomyopathy, atrial fibrillation, and hypercoagulability; and 6) Major findings from the PheCode enrichment analysis were replicated among AFR participants or the replication cohort (p_adj_ < 0.1 for PheCode sets derived from circulatory system from both cohorts).

Our phenome-wide analyses on RVs carriers with mostly heterozygous genotypes confirmed the pleiotropic and non-additive effects which were reported on homozygous carriers of PTVs, so-called “human knockouts” in a UK Biobank study^45^. We also confirmed that the *HTRA1* carriers had an increased risk for IS, particularly in the late-onset EUR participants. There is a conflict report between *in vitro* functional study^2,46^ and *in silicon*-based prediction^47^ about whether or not the *HTRA1*-p.Gln151Lys variant is deleterious. Most *HTRA1* carriers with EUR ancestry (103/120) were heterozygous for p.Gln151Lys, located at N-terminal domain with IGFBP and Kazal homology^46^.

According to the Kaplan-Meier analyses (**Supplementary Figure 9)**, p.Gln151Lys carries alone also had an increased risk for late-onset IS, suggesting its damaging effect. We argued that it is still valid to develop a knockout model to determine the functional effect of *HTRA1* IGFBP/Kazal domain within and beyond the cardiovascular system despite its *in vitro* null effect on protease activity^46^.

Our finding indicated that female carriers had a significantly increased risk for IS during menopause and the elderly stage than male carriers(Figure 4), suggesting there may have some cause for the incomplete penetrance and variable expressivity in male compared to females^9^, and it is important to recognize the sex differences in precision medicine for IS^48^.

Some of the findings identified in EUR participants were not replicated, likely due to their small effect sizes in the EUR, overrepresentation of participants of younger age, and some RVs being over- or under-represented in the replication cohort^10^. Given the extensive connection among PheCodes observed in the discovery EUR but not in the replication EUR (**Figure 2**), the disease trajectories for carriers in the discovery EUR could be more complicated than those observed in replication cohorts. This complexity may determine if the PheCode sets derived from one cohort show significant enrichment in the other cohort.

Various factors may account for the possibility of replication in an independent cohort. The validity of the results is determined by empirical variant curation (ACMG Clinvar annotation^49^) or machine learning algorithms (VEP annotation^50^ + additional approach^32^), which are used to prioritize functional variants. Using a comparative approach, we identified some discrepancies due to the variant annotation pipelines (**Supplementary Figure 3**). This discrepancy could result from the varied functional impact of each variant, their allele frequency and conservation, and their interaction with environmental factors (lifestyle, social determinants, and more) at the patient level.

### Observations and conclusions made from monogenic diseases

#### 1. RVs with large effects

The main driver for some variants with extremely low frequency is due to natural selection; however, the ‘reversal rare’ theory for protective alleles also exists^45^. Thus, the success of this study depends on the number of genuinely causal variants, the number of neutral variants or protective variants diluting signals, and whether the direction of effect is consistent within the gene. As we mentioned earlier, the additional filtering process for RVs would help enrich true deleterious RVs with higher penetrance. Type II error may occur due to including fewer RVs for some candidate genes.

#### 2. The omnigenic model – Core and peripheral genes

Most genes affect disease risk through highly connected gene regulatory/expression networks^51^. While RVs from the core genes demonstrate a larger effect size, peripheral genes exert their effects through interaction with core genes. For an omnigenic disease model, adding more ‘peripheral’ genes into the ‘core’ genes could lead to the diluted impacts with an increase in the false-positive results^10,51^. “The omnigenic model predicts that virtually any variant with regulatory effects in a given tissue is likely to have (weak) effects on all diseases that are modulated through that tissue”^51^. Instead of testing RVs individually, grouping variants with a similar function or belonging to genes within a similar functional network could boost the power of the association study. Pathway enrichment analysis of *COL4A1, NOTCH3*, and *HTRA1* coexpression networks in approximately 20 transcriptomic datasets from cardiovascular diseases showed ‘extracellular matrix organization’ as the top GO Biological Process terms (https://seek.princeton.edu/seek/). *COL4A1, NOTCH3*, and *HTRA1* are also highly correlated at the transcriptional level. In contrast, *CTC1* (T cell receptor signaling pathway), *GLA* (cellular response to lipopolysaccharide), and *TREX1*(cellular response to interferon) were mostly related to inflammation and immune response. Therefore, the overall effect from the seven candidate genes mainly reflected the disease profile due to the damage of the extracellular matrix or matrisome disfunction^52^.

#### 3. Pleiotropy and disease trajectory

Horizontal and vertical pleiotropy of monogenic variants results in diverged disease profile and trajectory. Extracerebral phenotypes are common in monogenic CSVD risk genes. The large-scale population-based phenome-wide longitudinal study of health outcomes could promote a better understanding of under-investigated disease trajectory beyond the brain^21^. Deep Phenotyping and Phenotypes conversion (stacking, orthogonal, deconvolution) enhance computing trajectories from wellness to disease for genetically predisposed individuals^53^. We must be cautious when the PTVs are enriched by the pipeline 2 (P2) selection; the latter may lead to patients with different disease trajectories or more patients with certain early-onset diseases compared to damaging missense variants enriched by P1 and P3 selections (**Supplementary Figure 3**). A similar observation of the rare or ultra-rare PTVs versus damaging missense variants across the phenotypic spectrum was also reported^54^. Genetic screening of patients with an early-onset of circulatory diseases for RVs of CSVD risk genes would prioritize individuals for continuous granular monitoring and promote personalized management.

### Study strength and limitation

This study had some strengths, such as leveraging rich longitudinal EHR, comparing three variants-annotation pipelines, and conducting PheCodes enrichment and sensitivity analyses to determine the sex- and age-dependent association. However, the study also has some limitations. First, the small sample size for participants reporting AFR ancestry and other ethnicities may prevent us from adequately estimating the effect size from those populations. Second, a potential underdiagnosis for carriers with mild phenotype is an inherent challenge of EHR-based observational study^55^. This would lead to an underestimation of the prevalence of genetically predisposed traits as the early sign or symptoms of advanced phenotypes (e.g., stroke or heart failure)^17^.

Pathogenic variants resulting in mild phenotypes may survive natural selection during reproductive age but increase the risk for mid- and late-onset diseases. Third, RVs identified by the three pipelines were limited to missense, splicing, and stop gain/lost sequence variants. Finally, the pleiotropic effect and varied effect sizes for each RV complicated the overall disease profiles and trajectories assessment. The varied expressivity of carriers could be due to an interplay between complex (polygenic) and Mendelian (monogenic) genetics^14^ for IS; the common disease is more complex than the implied “core genes” model^56^. Polygenic Risk Scores (PRS) derived from common variants could modify the outcome of patients with a monogenic disease^14,57,58^. How PRS modify the outcome of patients with RVs of monogenic risk genes will be the future focus.

## Conclusion

- Some common cardiovascular disease-related phenotypes (TIA, primary cardiomyopathy, atrial fibrillation, and hypercoagulability) were identified at an increased relative risk in RV carriers of seven CSVD risk genes.
- RVs from *COL4A1/2* or *NOTCH3* and *TREX1* were associated with IS in participants with AFR or EUR ancestry, respectively. Effect sizes are significantly larger in female participants. *HTRA1* carriers with EUR ancestry also showed a similar trend of association.
- Diseases from the circulatory system were significantly enriched in carriers with both EUR and AFR ancestry.
- Our findings support the concept of developing a gene panel of CSVD for population screening of patients with early-onset circulatory diseases. Under this clinical scenario, the diagnostic attribution through genetic testing is actionable. Whether the inclusion of CSVD genes on population-based genomic screening/ACMG secondary findings lists (which would be phenotype-agnostic) remains debatable.

## Data Availability

All data produced in the present study are available upon reasonable request to the authors. The codes are available at GitHub - TheDecodeLab/Trajectory-of-Monogenic-Diseases: Development of a platform to observe and analyses the trajectory of monogenic diseases.

## Glossary

ACMG: American College of Medical Genetics and Genomics
AFR: African ancestry
CI: confidence interval
CoxPH: Cox proportional-hazards model
CSVD: cerebral small vessel disease
EHR: Electronic Health Record
eQTL: expression quantitative trait loci
EUR: European ancestry
GO: Gene Ontology
GWAS: genome-wide association study
Hazards ratio: HR
HGMD: Human Gene Mutation Database
HWE: Hardy-Weinberg Equilibrium
ICD: International Classification of Disease
IS: IS
LAS: large-artery strokes
LD: linkage disequilibrium
MAF: minor allele frequency
ML: machine learning
OR: odds ratio
PSEA: PheCode Set Enrichment Analysis
PTV: protein truncation variants
RD: risk difference
RR: relative risk
RV: RV
PCA: Principal Component Analysis
PRE: percent relative effect
PRS: polygenic risk score
SNP: single nucleotide polymorphism
TIA: transient ischemic attack
VEP: Ensembl Variant Effect Predictor
VUS: a variant of uncertain significance.

