## Supplemental Figure 1-9 for "Race- and sex-specific disease trajectory in individuals with rare variants in seven cerebrovascular small vessel disease genes: a genotype-first population study"

**Supplementary Figure 1**. The prioritized rare variants from seven monogenic CSVD genes identified by three annotation pipelines were illustrated by UCSC genome browser. Their functional consequence as well as genotype counts were mapped to RefSeq sequence from NCBI, Pfam functional domain, ClinVar SNVs submitted interpretations and evidence, HGMD Variants, and OMIM Gene Phenotypes.

**Gene: HTRA1,**
**Synonyms: PRSS11, ARMD7, CARASIL, CADASIL2,**
**Phenotype: {Macular degeneration, age-related, neovascular type}, 3; {Macular degeneration, age-related, 7}, 3; CARASIL syndrome, AR, 3; Cerebral arteriopathy, autosomal dominant, with subcortical infarcts and leukoencephalopathy, type 2, AD, 3**

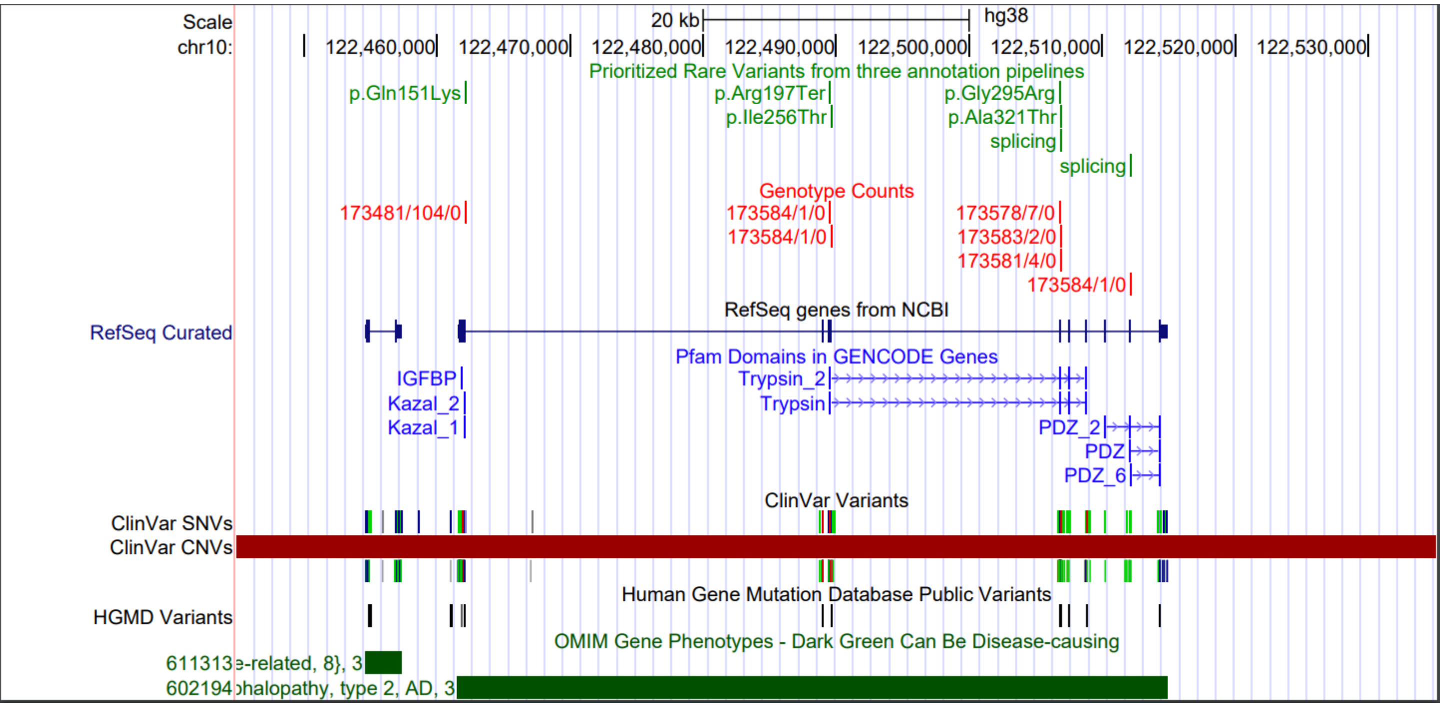


**Gene: CTC1,**
**Synonyms: CRMCC, C17orf68, AAF132,**
**Phenotype: Cerebroretinal microangiopathy with calcifications and cysts, AR, 3**

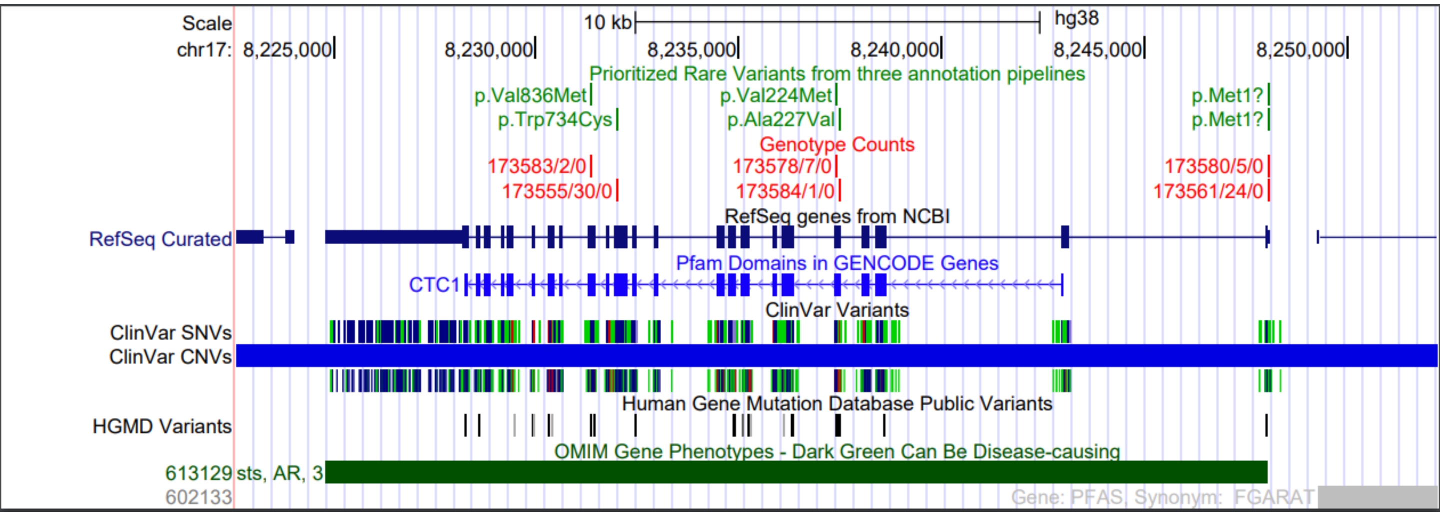

Supplementary Figure 1.

**Gene: COL4A1,**
**Synonyms: BSVD1, HANAC, ICH, BSVD, RATOR, PADMAL**
**Phenotype: ?Retinal arteries, tortuosity of, AD, 3; {Hemorrhage, intracerebral, susceptibility to }, 3; Angiopathy, hereditary with nephropathy, aneurysms, and muscle cramps, AD, 3; Microangiopathy and leukoencephalopathy, pontine, autosomal dominant, AD, 3; Brain small vessel disease with or without ocular anomalies, AD, 3**

**Gene: COL4A2,**
**Synonyms: BSVD2, ICH,**
**Phenotype: Brain small vessel disease 2, AD, 3; {Hemorrhage, intracerebral, susceptibility to}, 3**

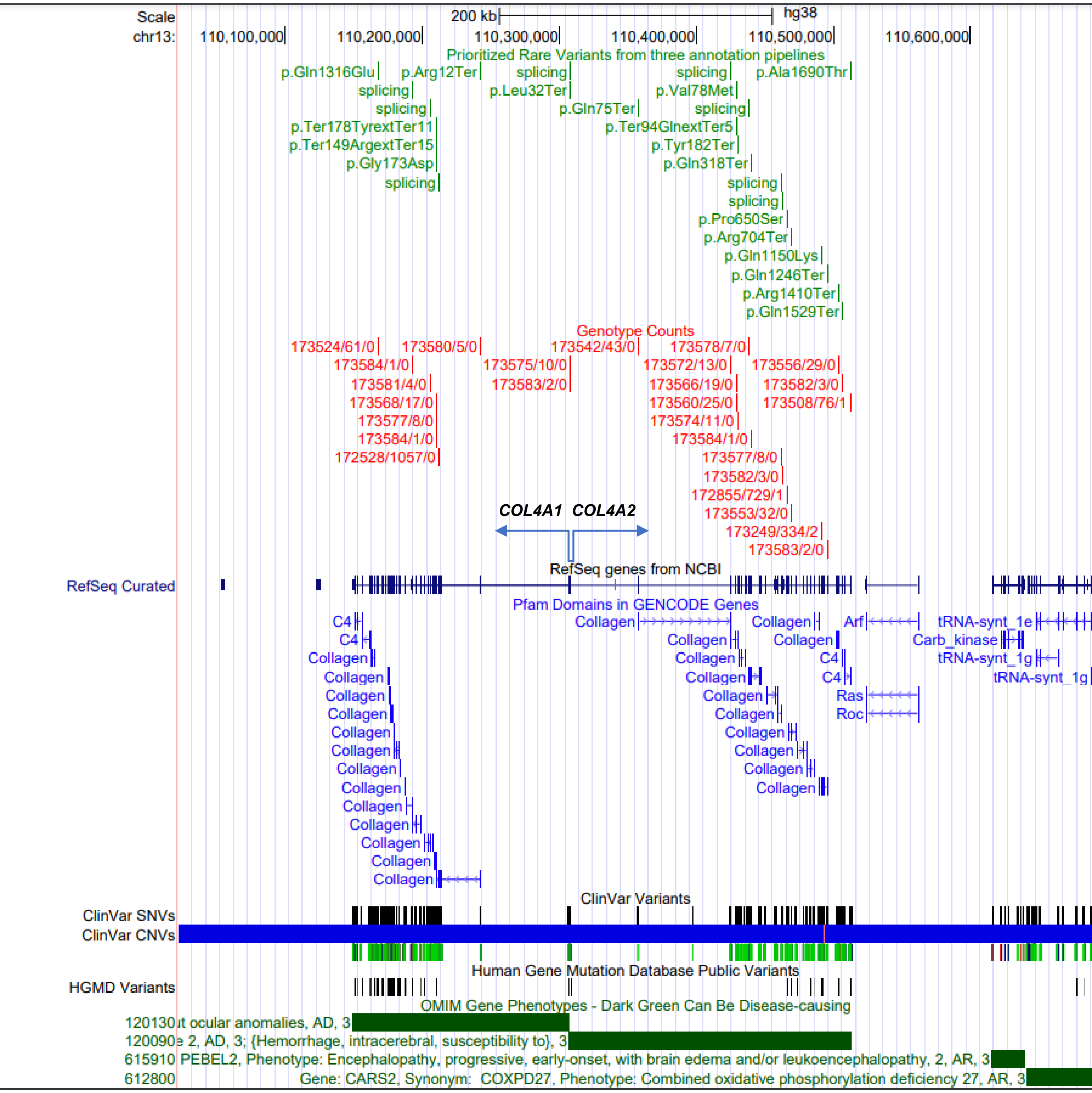

**Supplementary Figure 1.**

**Gene: NOTCH3,**
**Synonyms: CADASIL1, CASIL, IMF2, LMNS,**
**Phenotype: Lateral meningocele syndrome, AD, 3; ?Myofibromatosis, infantile 2, AD, 3; Cerebral arteriopathy with subcortical infarcts and leukoencephalopathy 1, AD, 3**

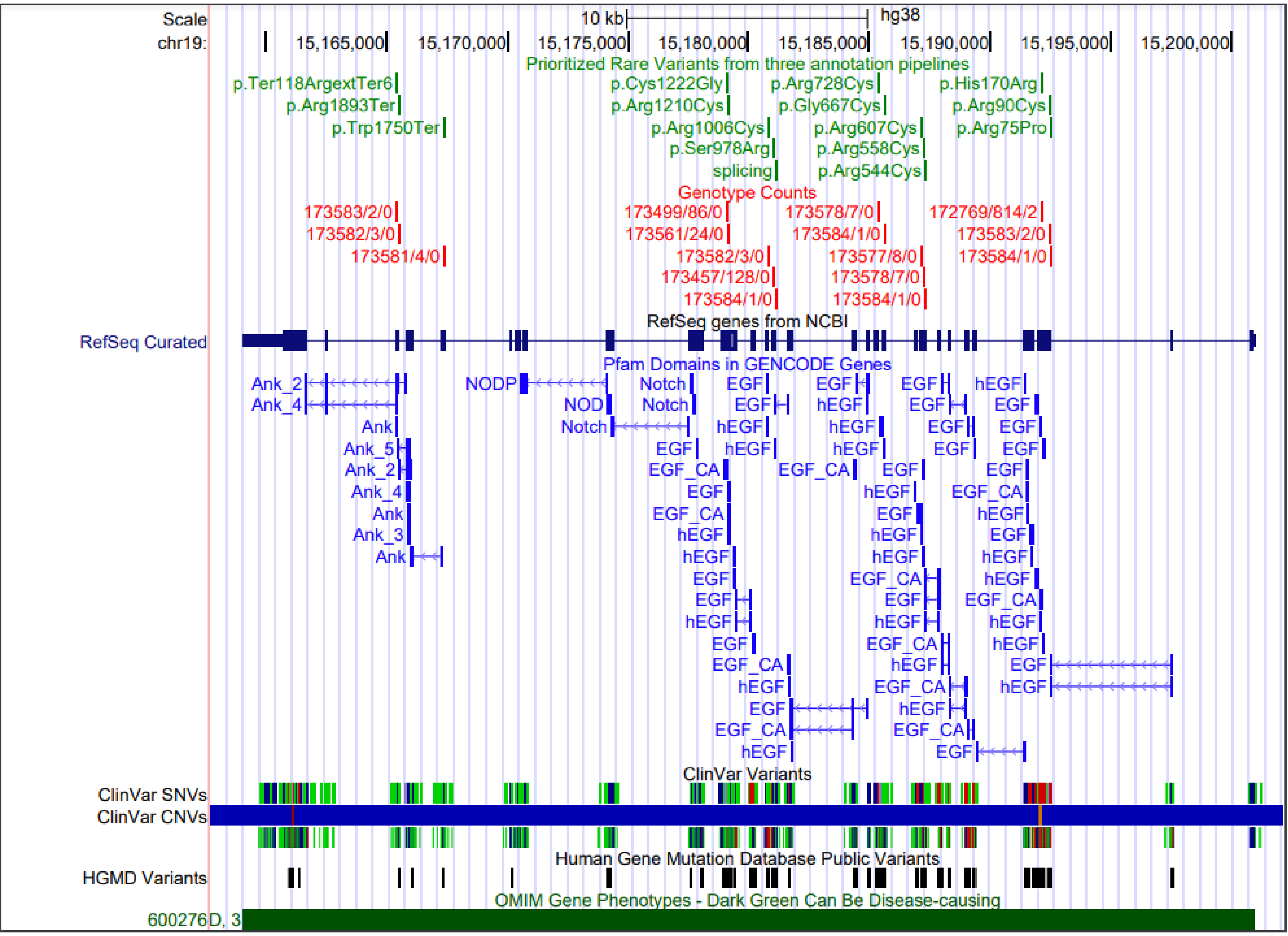

**Supplementary Figure 1.**

**Gene: TREX1,**
**Synonyms: AGS1, CRV, HERNS, RVCLS,**
**Phenotype: Vasculopathy, retinal, with cerebral leukoencephalopathy and systemic manifestations, AD, 3; Aicardi-Goutieres syndrome 1, dominant and recessive, AR/AD, 3; {Systemic lupus erythematosus, susceptibility to}, AD, 3; Chilblain lupus, AD, 3**

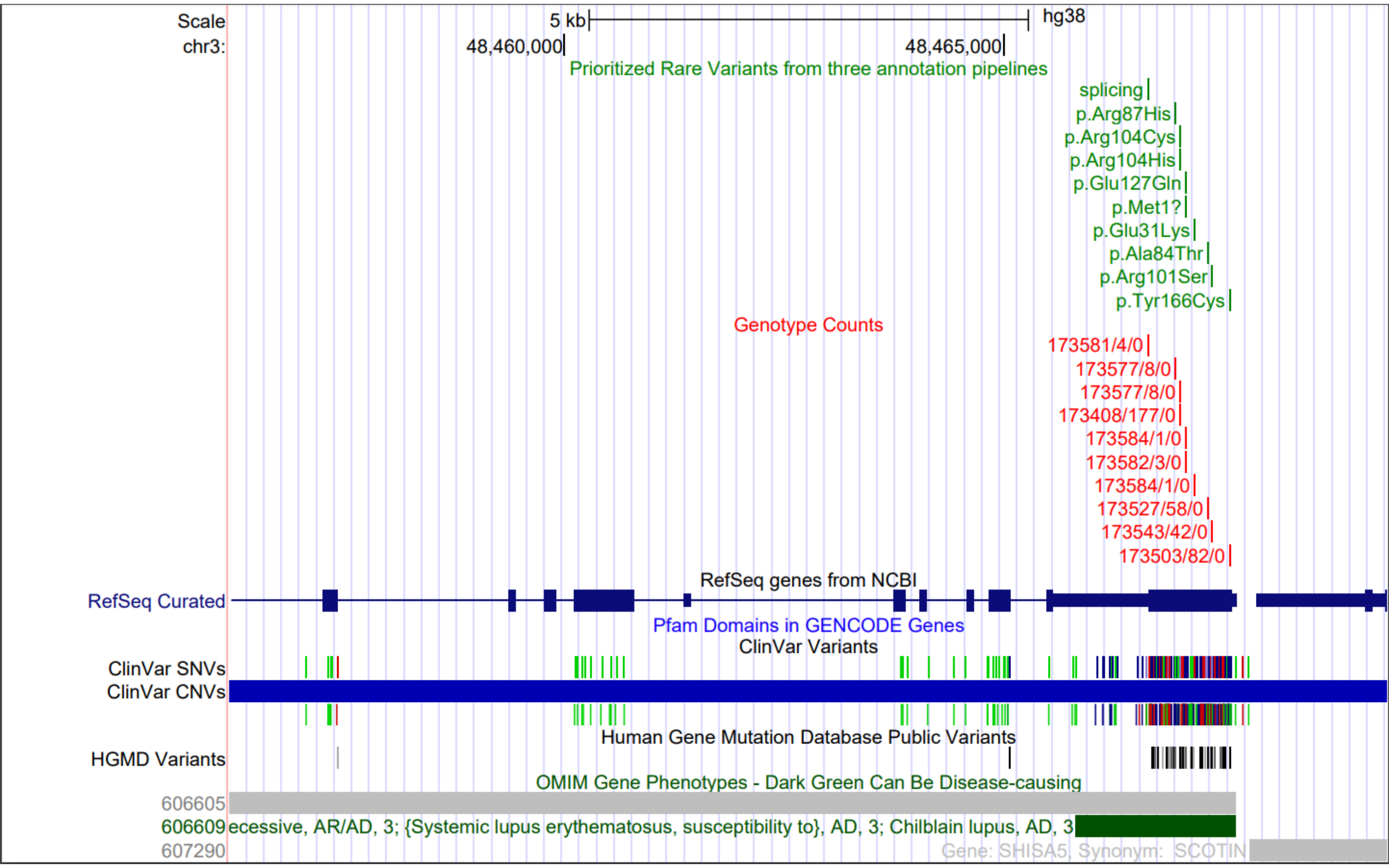


**Gene: GLA,**
**Phenotype: Fabry disease, cardiac variant, XL, 3; Fabry disease, XL, 3**

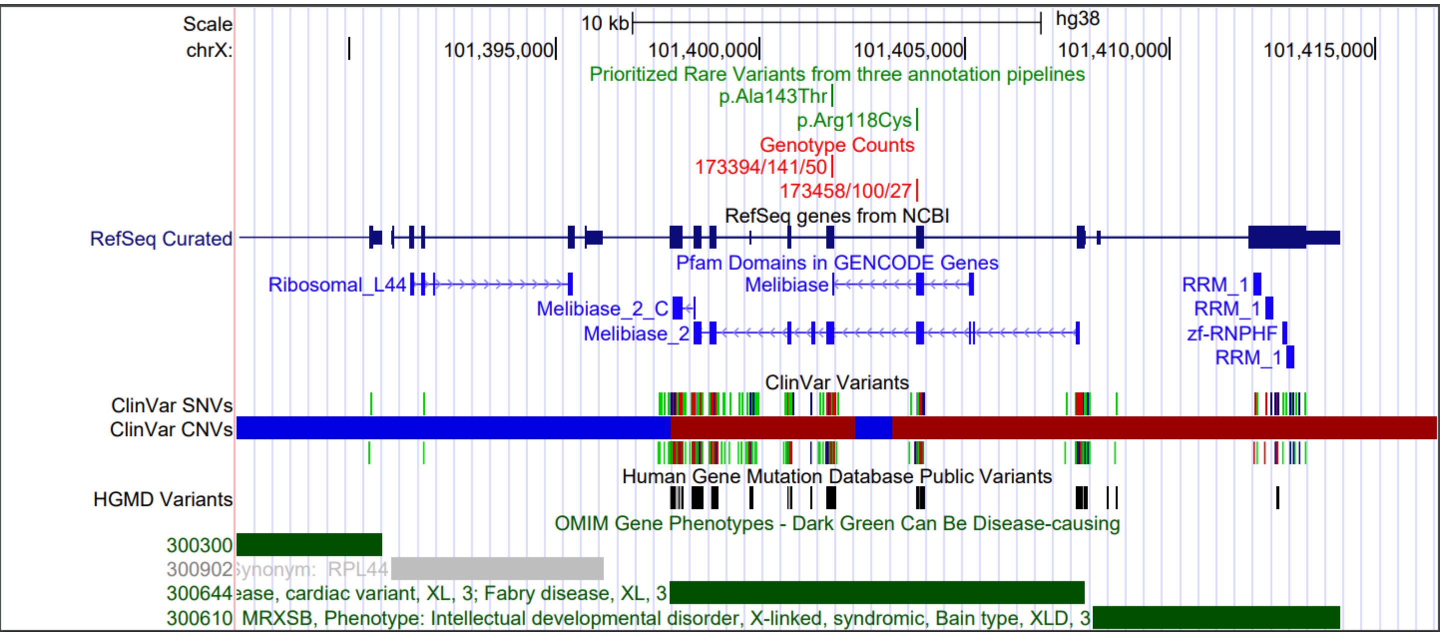

**Supplementary Figure 2. LOVE plot to display the improvement in standardized mean differences after propensity score matching (PSM) for index age, race, and sex.**

Age, race, and sex were identified as confounding factors for the association between rare variants and phenotypes. They were selected as covariates in a Logistic Regression model to create propensity scores (R MatchIt package). We chose “nearest neighbor matching without replacement” to create a more balanced ratio (1:2) for carriers and noncarriers. The love plot (R cobalt package) demonstrated how well the improvement of standardized mean differences deviated from zero for each covariate before and after PSM in Discovery (left) and Replication (right) samples after PSM at 1:2 ratio.

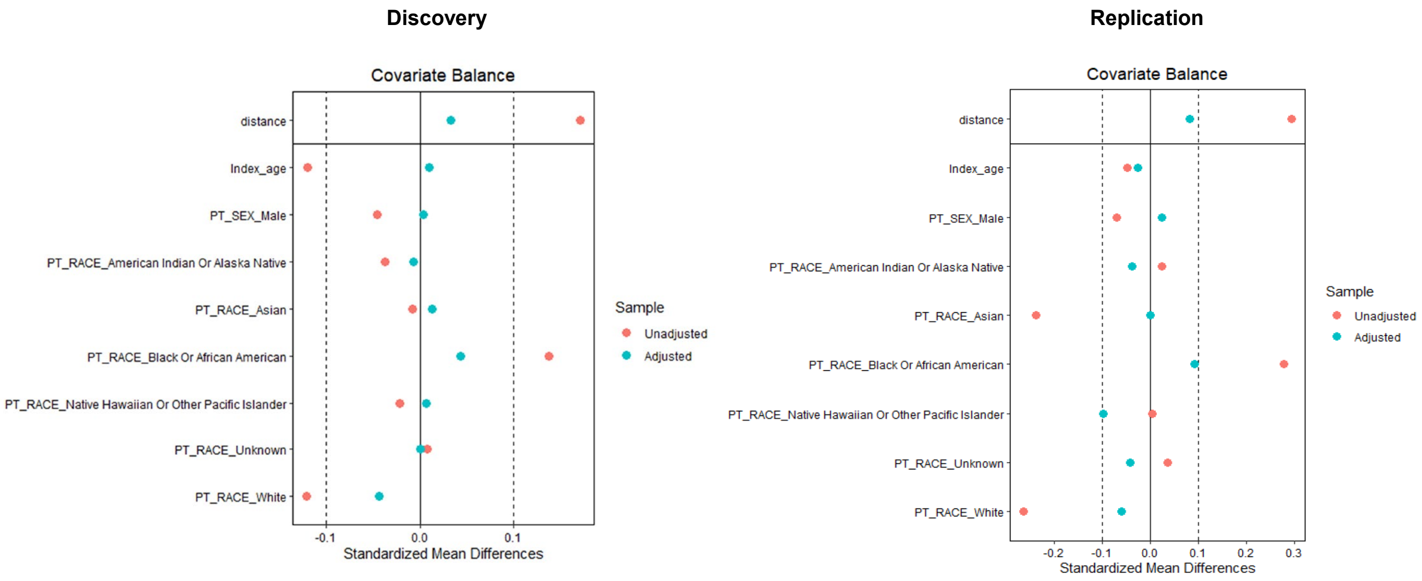

**Supplementary Figure 3. Comparing the rare variants and the corresponding carriers identified from three pipelines in both discovery and replication cohorts.** Venn Diagram showed the distribution of rare variants prioritized by three pipelines. We chose 20 or 22 PheCodes from circulatory system for both cohorts, respectively. The description of PheCodes were listed in the corresponding tables. The goal is to determine which pipeline identify more patients for the specific PheCode. Percent Relative Effect (PRE) derived from three pipelines as well as combination of intersects between pipelines was considered as the metric to evaluate the performance. The functional consequence annotated by VEP was list in the table. Some variants may belong to two more categories depending on the reported transcripts collected in the Ensembl database as shown in Supplementary Table 1.

### Discovery (≤65yrs)

Intersect : (P1∩P2) U (P1∩P3) U (P2∩P3)

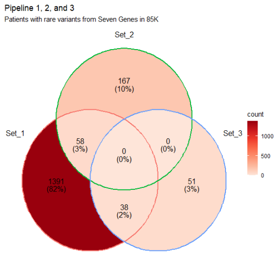


| phecode | phecode_str |
|---|---|
| 395 | Heart valve disorders |
| 395.1 | Nonrheumatic mitral valve disorders |
| 395.2 | Nonrheumatic aortic valve disorders |
| 396 | Abnormal heart sounds |
| 401.3 | Other hypertensive complications |
| 420 | Carditis |
| 425 | Cardiomyopathy |
| 425.1 | Primary/intrinsic cardiomyopathies |
| 426.7 | Abnormal electrocardiogram [ECG] [EKG] |
| 429.3 | Symptoms involving cardiovascular system |
| 433 | Cerebrovascular disease |
| 433.1 | Occlusion and stenosis of precerebral arteries |
| 433.12 | Cerebral atherosclerosis |
| 433.2 | Occlusion of cerebral arteries |
| 433.21 | Cerebral artery occlusion, with cerebral infarction |
| 433.3 | Cerebral ischemia |
| 433.31 | Transient cerebral ischemia |
| 443.1 | Raynaud's syndrome |
| 452 | Other venous embolism and thrombosis |
| 452.2 | Deep vein thrombosis [DVT] |

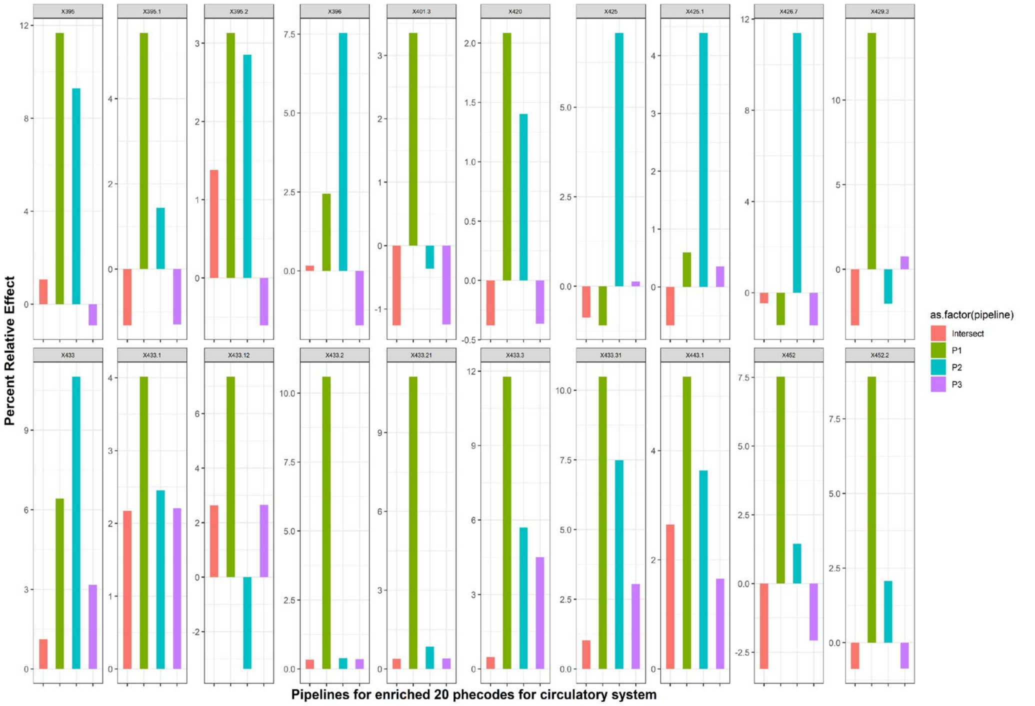


### Replication (≤65yrs)

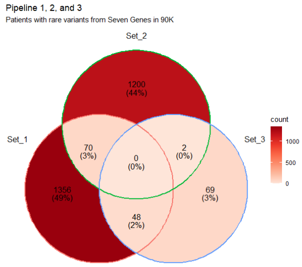


| phecode | phecode_str |
|---|---|
| 401.2 | Hypertensive heart and/or renal disease |
| 401.21 | Hypertensive heart disease |
| 411.2 | Myocardial infarction |
| 411.4 | Coronary atherosclerosis |
| 414 | Other forms of chronic heart disease |
| 415 | Pulmonary heart disease |
| 425 | Cardiomyopathy |
| 425.1 | Primary/intrinsic cardiomyopathies |
| 426 | Cardiac conduction disorders |
| 426.7 | Abnormal electrocardiogram [ECG] [EKG] |
| 426.9 | Cardiac pacemaker/device in situ |
| 427.2 | Atrial fibrillation and flutter |
| 427.21 | Atrial fibrillation |
| 427.7 | Tachycardia NOS |
| 428.1 | Congestive heart failure (CHF) NOS |
| 428.3 | Heart failure with reduced EF [Systolic or combined heart failure] |
| 433 | Cerebrovascular disease |
| 433.3 | Cerebral ischemia |
| 433.31 | Transient cerebral ischemia |
| 440 | Atherosclerosis |
| 443.9 | Peripheral vascular disease, unspecified |
| 447 | Other disorders of arteries and arterioles |

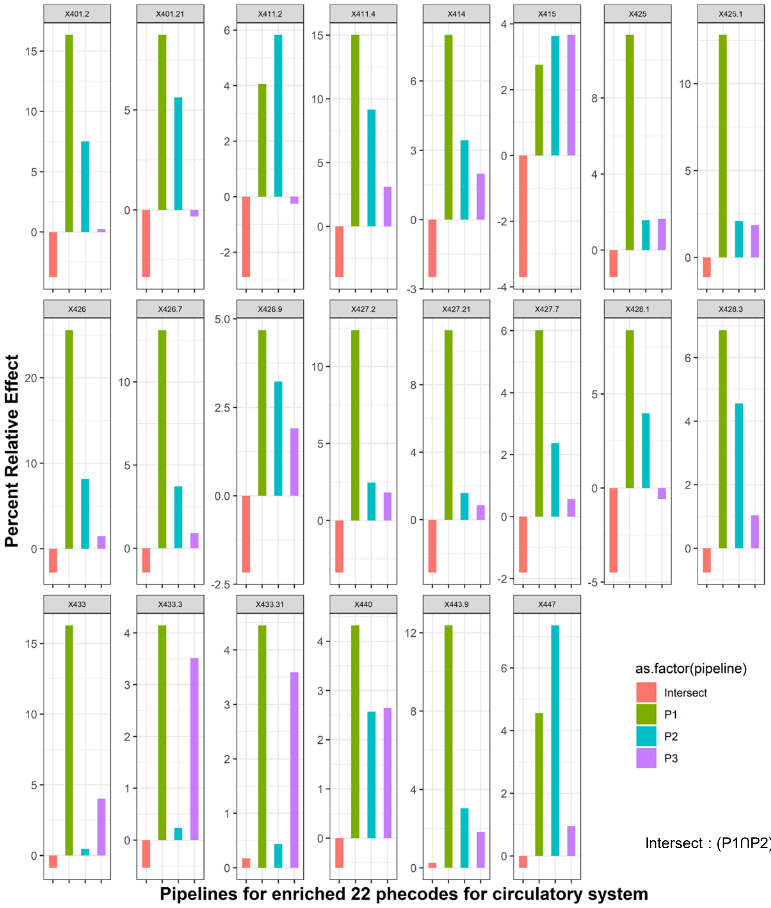


Intersect : (P1∩P2) U (P1∩P3) U (P2∩P3)

| VEP annotation category | Number |
|---|---|
| intergenic_variant | |
| feature_truncation | |
| regulatory_region_variant | |
| feature_elongation | |
| regulatory_region_amplification | |
| regulatory_region_ablation | |
| TF_binding_site_variant | |
| TFBS_amplification | |
| TFBS_ablation | |
| downstream_gene_variant | |
| upstream_gene_variant | |
| non_coding_transcript_variant | |
| NMD_transcript_variant | 8 |
| intron_variant | |
| non_coding_transcript_exon_variant | |
| 3_prime_UTR_variant | |
| 5_prime_UTR_variant | |
| mature_miRNA_variant | |
| coding_sequence_variant | |
| synonymous_variant | 1 |
| stop_retained_variant | |
| incomplete_terminal_codon_variant | |
| splice_region_variant | |
| protein_altering_variant | |
| missense_variant | 38 |
| inframe_deletion | |
| inframe_insertion | |
| transcript_amplification | |
| start_lost | 3 |
| stop_lost | 4 |
| frameshift_variant | |
| stop_gained | 12 |
| splice_donor_variant | 5 |
| splice_acceptor_variant | 7 |
| transcript_ablation | |

**Supplementary Figure 4. Circo plots with interlinks of the top 50 common diseases of which rare variants from six CSVD risk genes having increase relative risk during lifetime observation.** A circo plot was created based on the top 50 PheCodes ranked by percent relative effect (PRE) and clustered by the processes. The links represent the significant association between two PheCodes in rare variant carriers using Fisher's exact test after correction for multiple testing ($p < 0.001$ given 50 phecodes being tested, $1/(50*49)/2) \approx 0.001$). Significant association for adjacent PheCodes were labeled with spike but not linked. Y axis represent PRE; The area of the dot was proportional to the significant association between mutation and the corresponding disease according to Fisher's exact test. Diseases from circulatory system were overrepresent in rare variant carriers in both cohorts with AFR ancestry but not with EUR ancestry. Categories with the number of PheCodes >= 2 were listed.

**Discovery** **Replication**

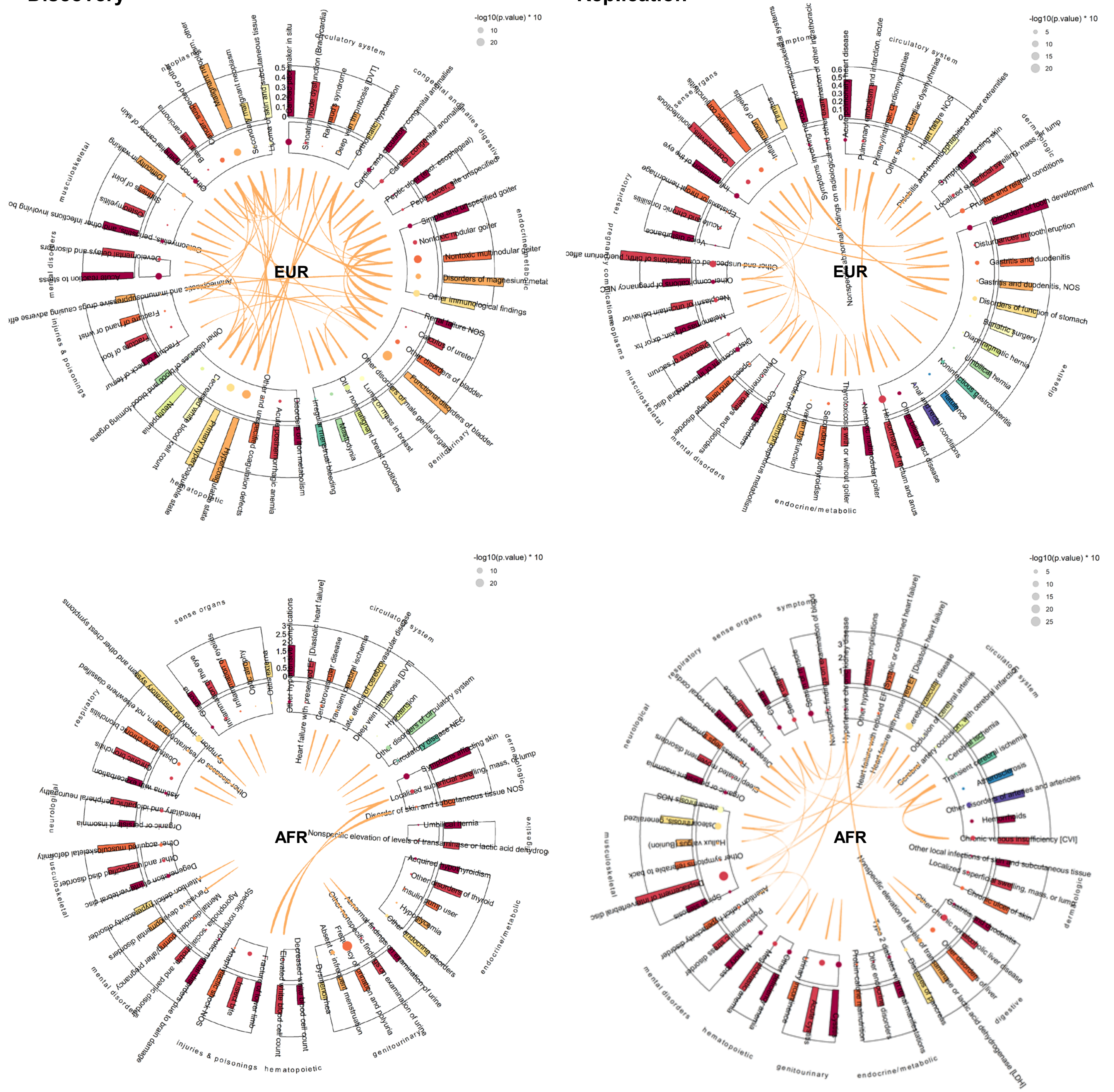

**Supplementary Figure 5. For individual CSVD risk gene, PheCodes from circulatory system were overrepresented in the top 50 common diseases with increased relative risk in both or one of cohorts.** We compared rare variant carriers from individual gene to noncarriers for all six CSVD genes. Patients with age of onset after 65yrs considered null for a specific disease. Categories with the number of PheCodes >= 2 were listed.

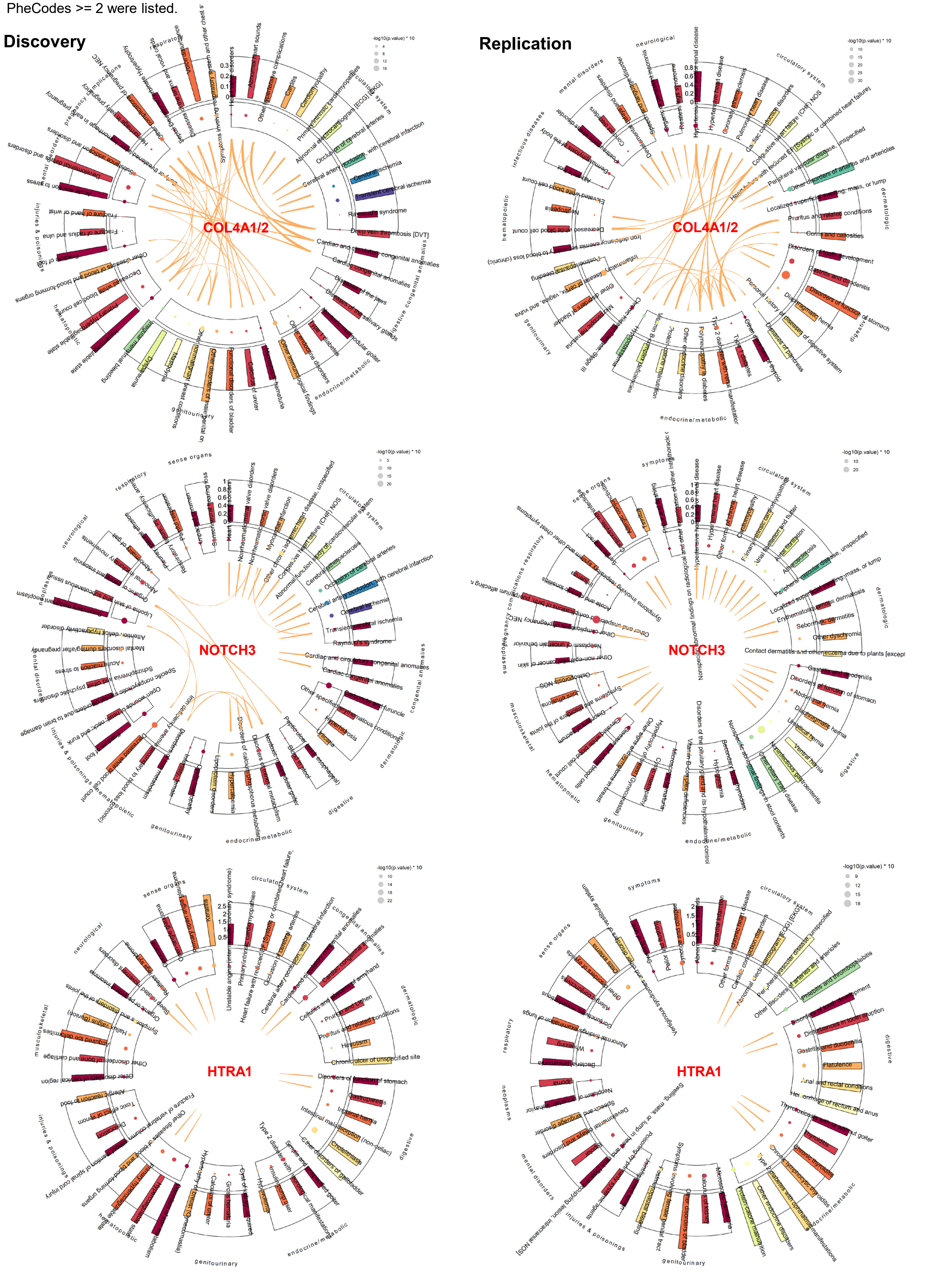

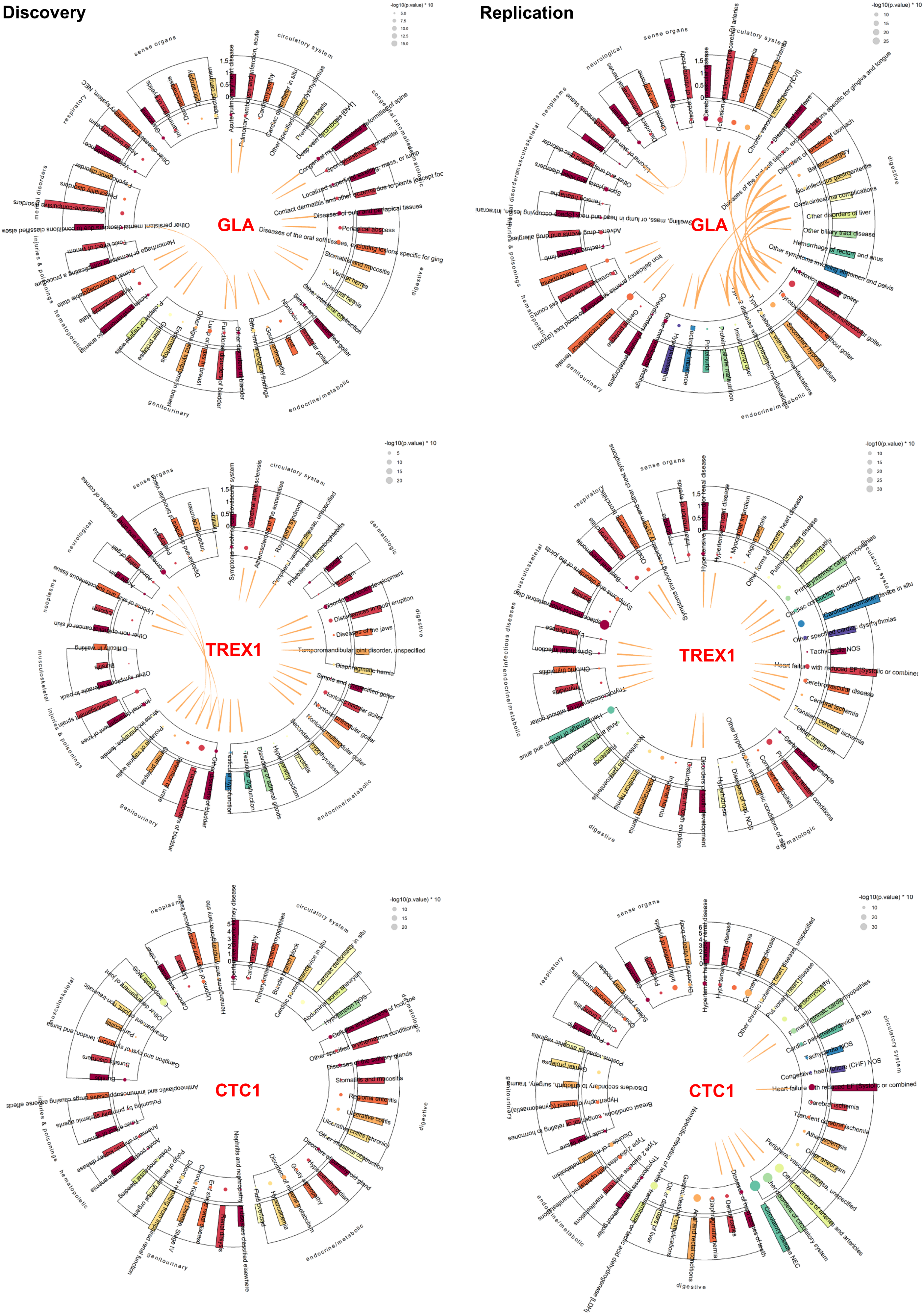

Discovery
Replication
GLA
TREX1
CTC1
-log10(p.value) * 10
sense organs
circulatory system
respiratory
mental disorders
injuries & poisonings
hematopoietic
genitourinary
endocrine/metabolic
digestive
congenital anomalies
dermatologic
neurological
neoplasms
musculoskeletal
infectious diseases

**Supplementary Figure 7. Kaplan-Meier estimated the cumulative probability of hypercoagulability and primary cardiomyopathy for carriers comparing to matched noncarriers. A gene-dependent analysis in EUR patients.**

*Cumulative probability for 85yrs*

Disease = hypercoagulability 3 phecodes

A.

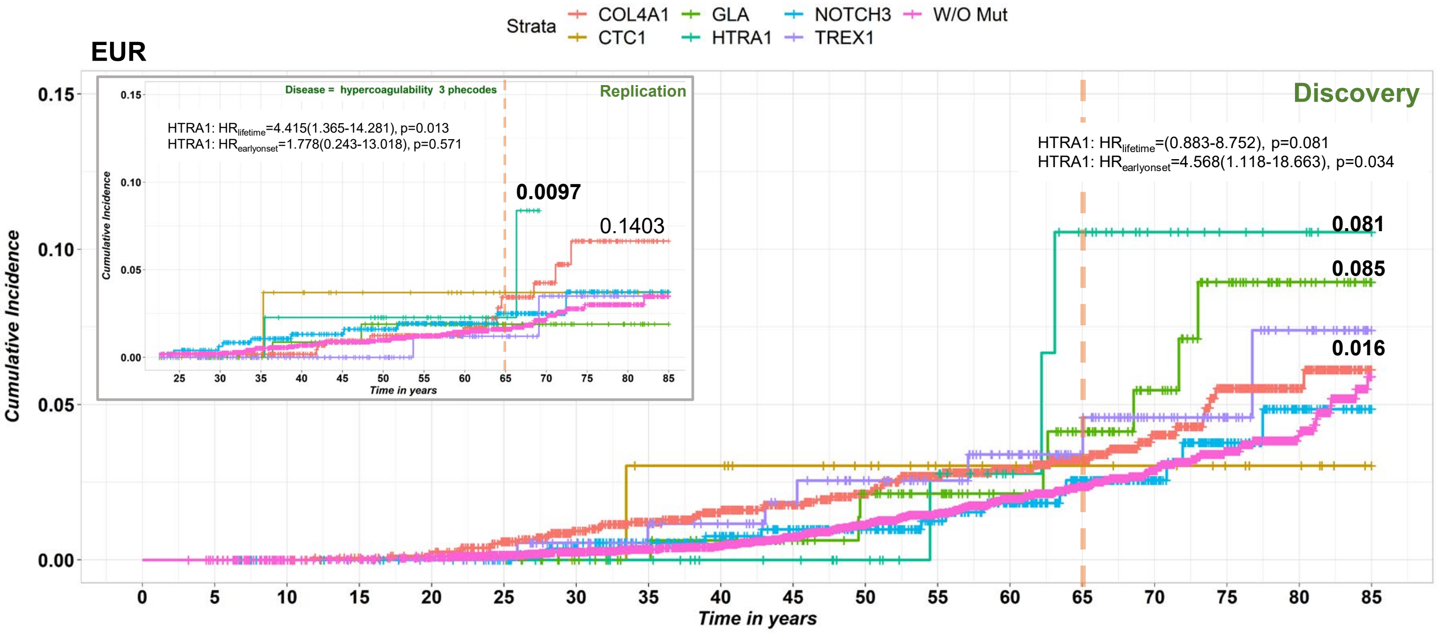


*Cumulative probability for 85yrs*

Disease = Primary/intrinsic cardiomyopathies (425.1)

B.

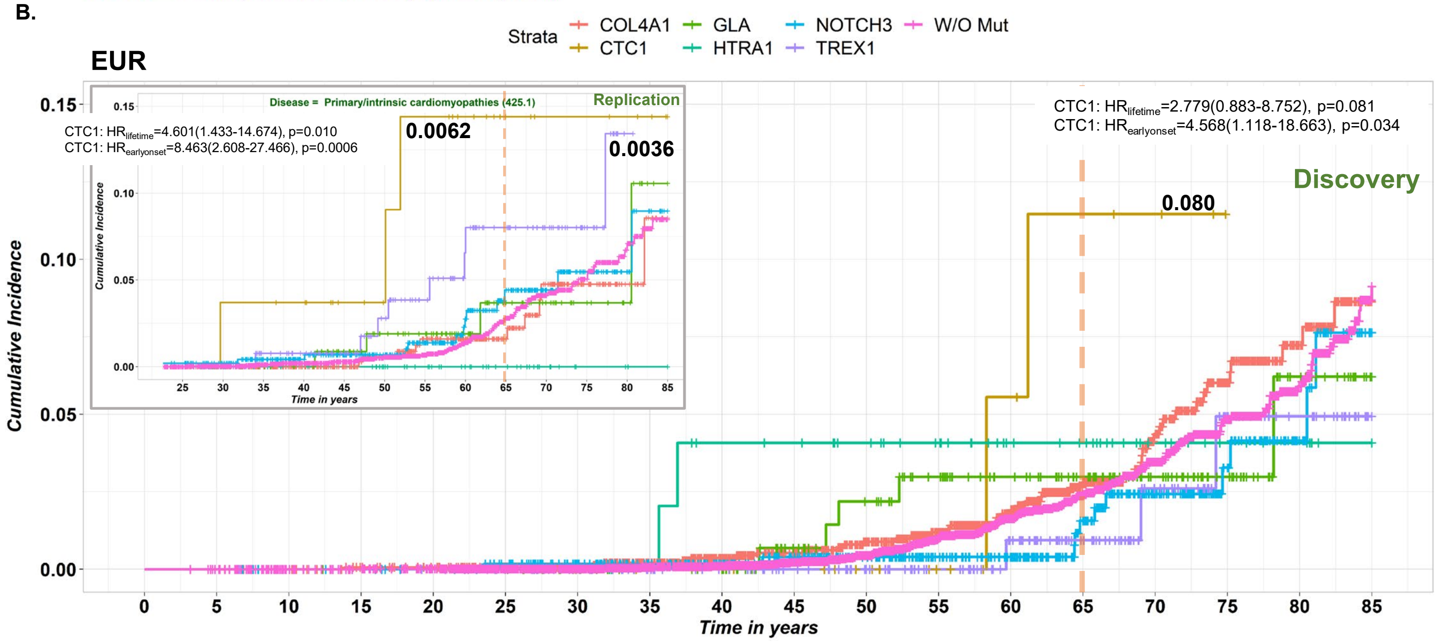

**Supplementary Figure 8. PheCode Set Enrichment Analysis for validation across cohort and/or across race using the PheCode set derived from the circulatory system as examples.**

**A**. Summary of the PheCode set enrichment analysis and validation across cohort and/or across race; **B, C, and D** represent the process to obtain the PheCode set from each subcohort (Discovery AFR, Replication EUR, and Replication AFR, respectively) with the highest enrichment score for each disease category.

**A.**

| Rank_List | PheCode_set_List | NES | pval | padj |
|---|---|---|---|---|
| Discovery (EUR) | Discovery (AFR) | 0.77 | 8.40E-01 | 9.60E-01 |
| Discovery (AFR) | Discovery (EUR) | 1.61 | 1.90E-02 | 1.20E-01 |
| Discovery (EUR) | Replication (EUR) | 0.76 | 8.70E-01 | 9.70E-01 |
| Discovery (AFR) | Replication (AFR) | 1.75 | 3.00E-03 | 5.10E-02 |
| Replication (EUR) | Replication (AFR) | 1.89 | 9.80E-05 | 1.60E-03 |
| Replication (AFR) | Replication (EUR) | 1.98 | 5.00E-05 | 8.10E-04 |
| Replication (EUR) | Discovery (EUR) | 1.12 | 3.20E-01 | 9.50E-01 |
| Replication (AFR) | Discovery (AFR) | 1.72 | 4.90E-03 | 8.30E-02 |
| Discovery (EUR) | Discovery (EUR) | 2.95 | 1.70E-11 | 2.80E-10 |
| Discovery (AFR) | Discovery (AFR) | 2.43 | 1.60E-10 | 2.70E-09 |
| Replication (EUR) | Replication (EUR) | 2.22 | 1.10E-08 | 1.90E-07 |
| Replication (AFR) | Replication (AFR) | 2.36 | 5.00E-09 | 8.50E-08 |
| Discovery (EUR) | Replication (AFR) | 1.69 | 9.60E-03 | 7.70E-02 |
| Discovery (AFR) | Replication (EUR) | 1.72 | 6.70E-03 | 5.40E-02 |
| Replication (EUR) | Discovery (AFR) | 1.63 | 2.20E-02 | 1.80E-01 |
| Replication (AFR) | Discovery (EUR) | 1.77 | 2.30E-03 | 3.10E-02 |

**B. Discovery(AFR)**

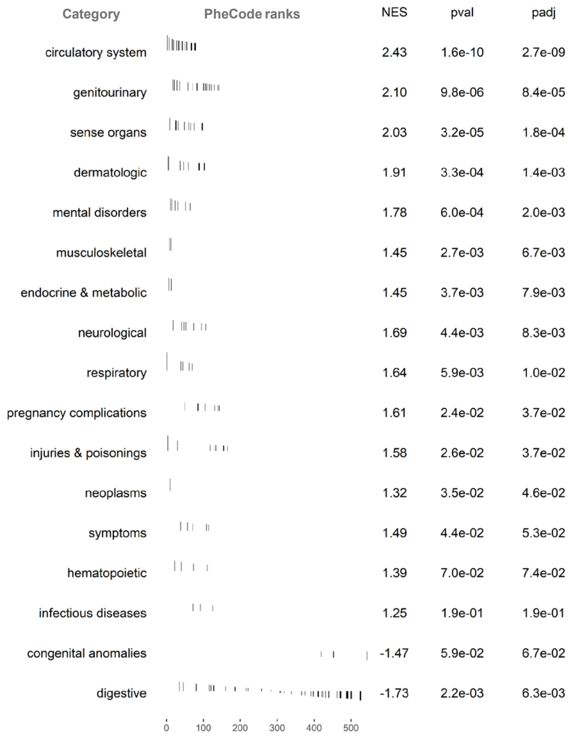


| Cut_point | Enrichment_score_max | Category | TotalNum | Num |
|---|---|---|---|---|
| 125 | 0.249 | infectious diseases | 8 | 3 |
| 10 | 0.627 | neoplasms | 4 | 1 |
| 13 | 0.131 | endocrine & metabolic | 64 | 2 |
| 110 | 0.423 | hematopoietic | 14 | 4 |
| 64 | 0.374 | mental disorders | 33 | 6 |
| 106 | 0.443 | neurological | 23 | 7 |
| 98 | 0.358 | sense organs | 35 | 12 |
| 79 | 0.474 | circulatory system | 66 | 20 |
| 69 | 0.302 | respiratory | 36 | 5 |
| 526 | 0.037 | digestive | 53 | 53 |
| 142 | 0.267 | genitourinary | 68 | 20 |
| 143 | 0.316 | pregnancy complications | 16 | 8 |
| 103 | 0.407 | dermatologic | 32 | 10 |
| 12 | 0.162 | musculoskeletal | 45 | 2 |
| 543 | 0.002 | congenital anomalies | 4 | 4 |
| 114 | 0.167 | symptoms | 25 | 5 |
| 165 | 0.381 | injuries & poisonings | 18 | 9 |

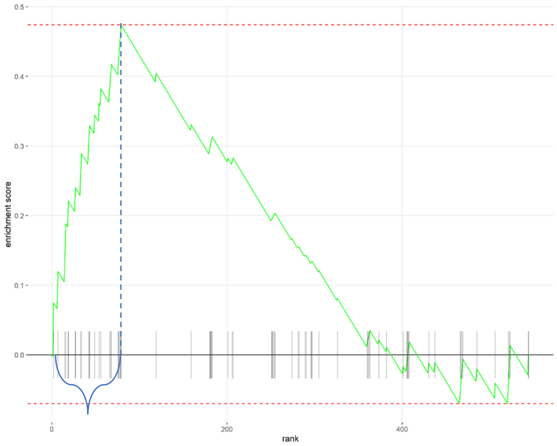


circulatory system ("396" "401.3" "411.3" "411.8" "425" "425.1" "426.9" "427.2" "427.21" "428.1" "428.2" "428.3" "429.2" "433" "433.3" "433.31" "443.9" "452.2" "459" "459.9"

### C. Replication(EUR)

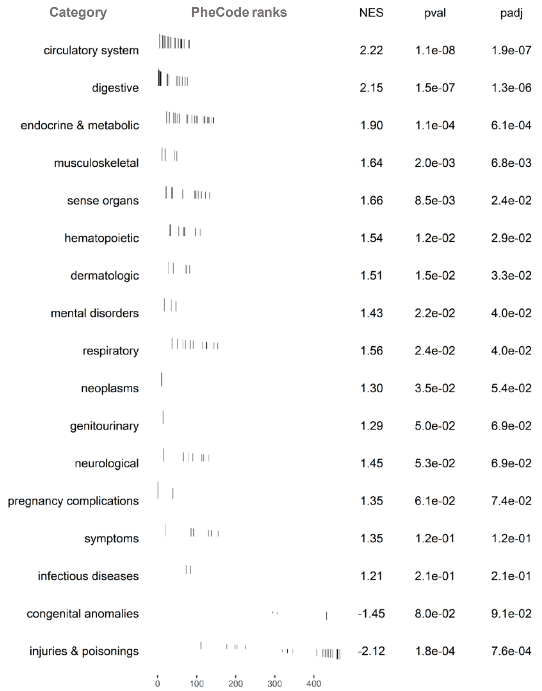


| Cut_point | Enrichment_score_max | Category | TotalNum | Num |
|---|---|---|---|---|
| 84 | 0.404 | infectious diseases | 8 | 2 |
| 10 | 0.319 | neoplasms | 10 | 1 |
| 143 | 0.239 | endocrine & metabolic | 58 | 21 |
| 109 | 0.552 | hematopoietic | 13 | 7 |
| 47 | 0.256 | mental disorders | 27 | 3 |
| 131 | 0.357 | neurological | 20 | 7 |
| 133 | 0.433 | sense organs | 26 | 11 |
| 81 | 0.517 | circulatory system | 52 | 20 |
| 154 | 0.342 | respiratory | 33 | 11 |
| 76 | 0.599 | digestive | 43 | 17 |
| 14 | 0.043 | genitourinary | 63 | 1 |
| 39 | 0.404 | pregnancy complications | 14 | 2 |
| 82 | 0.168 | dermatologic | 34 | 5 |
| 49 | 0.318 | musculoskeletal | 33 | 5 |
| 432 | 0.105 | congenital anomalies | 4 | 4 |
| 155 | 0.215 | symptoms | 24 | 7 |
| 466 | 0.035 | injuries & poisonings | 20 | 20 |

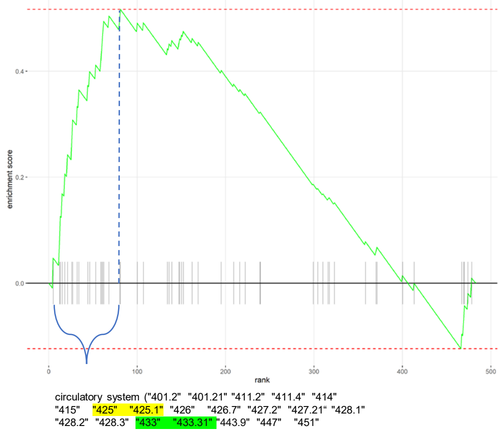


### D. Replication(AFR)

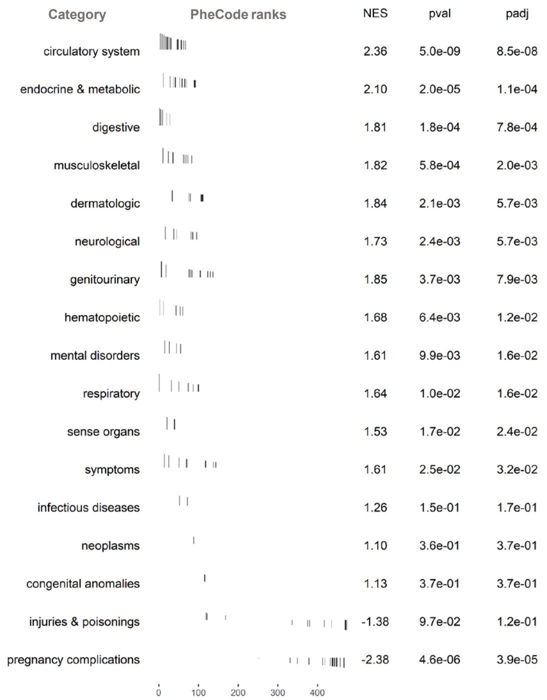


| Cut_point | Enrichment_score_max | Category | TotalNum | Num |
|---|---|---|---|---|
| 72 | 0.365 | infectious diseases | 7 | 2 |
| 88 | 0.062 | neoplasms | 5 | 1 |
| 92 | 0.393 | endocrine & metabolic | 57 | 15 |
| 60 | 0.511 | hematopoietic | 19 | 5 |
| 55 | 0.213 | mental disorders | 31 | 4 |
| 96 | 0.479 | neurological | 20 | 8 |
| 40 | 0.157 | sense organs | 31 | 3 |
| 67 | 0.566 | circulatory system | 48 | 18 |
| 100 | 0.350 | respiratory | 35 | 7 |
| 28 | 0.387 | digestive | 41 | 5 |
| 137 | 0.252 | genitourinary | 52 | 15 |
| 467 | 0.030 | pregnancy complications | 19 | 19 |
| 112 | 0.269 | dermatologic | 34 | 11 |
| 83 | 0.414 | musculoskeletal | 36 | 9 |
| 116 | 0.428 | congenital anomalies | 5 | 2 |
| 144 | 0.381 | symptoms | 24 | 10 |
| 0 | 0.000 | injuries & poisonings | 17 | 12 |
| 481 | 0.000 | injuries & poisonings | 17 | 12 |
| 482 | 0.000 | injuries & poisonings | 17 | 12 |

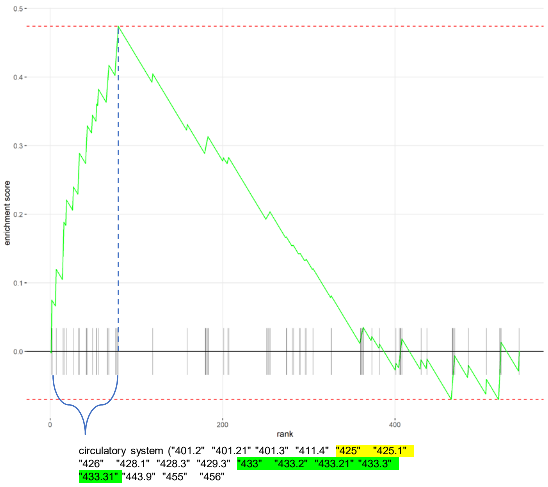

**Supplementary Figure 9. Kaplan-Meier analyses compare the cumulative probability of ischemic cerebrovascular disease for all *HTRA1* RV carriers versus *HTRA1* p.Gln151Lys carriers alone.** We combined both discovery and replication cohorts and only showed the results for patients with EUR ancestry **A.** p.Gln151Lys and other rare variants carriers; B. p.Gln151Lys carriers only.

**A.**

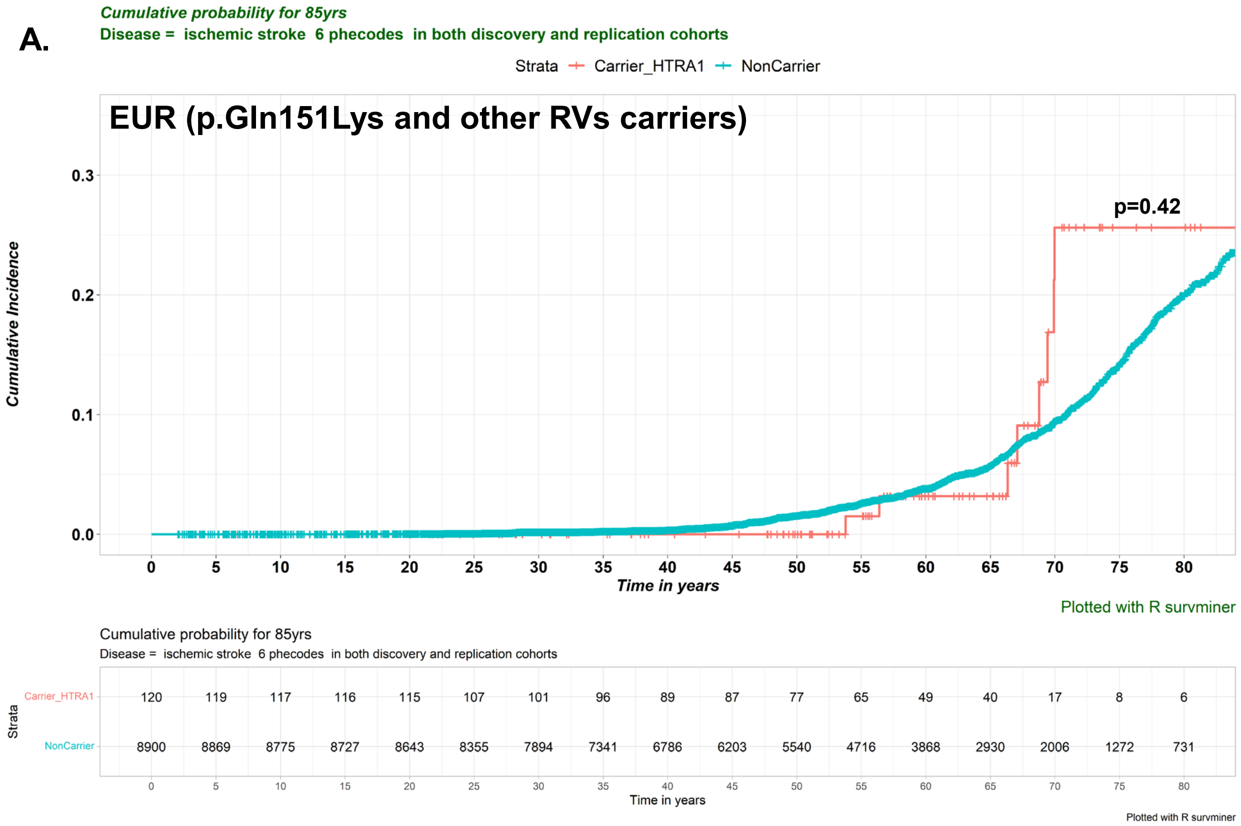


**B.**

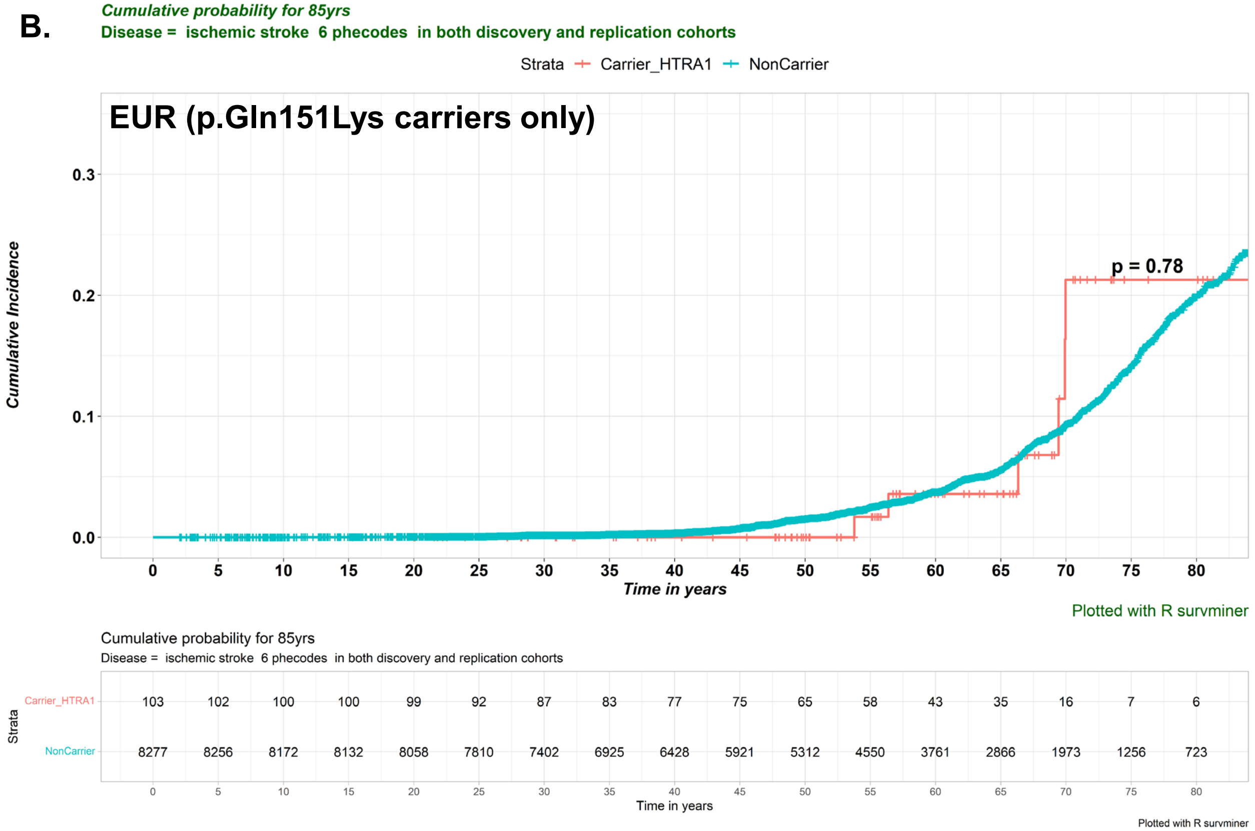
